# Direct and INdirect effects analysis of Genetic lOci (DINGO): A software package to increase the power of locus discovery in GWAS meta-analyses of perinatal phenotypes and traits influenced by indirect genetic effects

**DOI:** 10.1101/2023.08.22.23294446

**Authors:** Liang-Dar Hwang, Gabriel Cuellar-Partida, Loic Yengo, Jian Zeng, Robin N. Beaumont, Rachel M. Freathy, Gunn-Helen Moen, Nicole M. Warrington, David M. Evans

**Author notes:** Correspondence to Prof David M Evans, Institute for Molecular Bioscience, University of Queensland, Brisbane, Australia.

## Abstract

Perinatal traits are influenced by genetic variants from both fetal and maternal genomes. Genome-wide association studies (GWAS) of these phenotypes have typically involved separate fetal and maternal scans, however, this approach may be inefficient as it does not utilize the information shared across the individual GWAS. In this manuscript we investigate the performance of three strategies to detect loci in maternal and fetal GWAS of the same trait: (i) the traditional strategy of analysing maternal and fetal GWAS separately; (ii) a novel two degree of freedom test which combines information from maternal and fetal GWAS; and (iii) a novel one degree of freedom test where signals from maternal and fetal GWAS are meta-analysed together conditional on the estimated sample overlap. We demonstrate through a combination of analytical formulae and data simulation that the optimal strategy depends on the extent of sample overlap/relatedness between the maternal and fetal GWAS, the correlation between own and offspring phenotypes, whether loci jointly exhibit fetal and maternal effects, and if so, whether these effects are directionally concordant. We apply our methods to summary results statistics from a recent GWAS meta-analysis of birth weight from deCODE, the UK Biobank and the Early Growth Genetics (EGG) consortium. Both the two degree of freedom (213 loci) and meta-analytic approach (226 loci) dramatically increase the number of robustly associated genetic loci for birth weight relative to separately analysing the scans (183 loci). Our best strategy identifies an additional 62 novel loci compared to the most recent published meta-analysis of birth weight and implicates both known and new biological pathways in the aetiology of the trait. We implement our methods in the online DINGO (**D**irect and **IN**direct effects analysis of **G**enetic l**O**ci) software package, which allows users to perform one and/or two degree of freedom tests easily and computationally efficiently across the genome. We conclude that whilst the novel two degree of freedom test may be particularly useful for the analysis of certain perinatal phenotypes where many loci exhibit discordant maternal and fetal genetic effects, for most phenotypes, a simple meta-analytic strategy is likely to perform best, particularly in situations where maternal and fetal GWAS only partially overlap.

## Introduction

Perinatal traits like birth weight are influenced by genetic variants from both offspring and maternal genomes ^1-6^. Historically, genome-wide associations studies (GWAS) of these traits have involved separate analyses of offspring and maternal genomes i.e. a “fetal GWAS” where an individual’s own phenotype is regressed on their own genotype, and a separate “maternal GWAS” where offspring phenotype is regressed on maternal genotype. Genetic variants showing genome-wide significant associations in either scan are then followed up using conditional association analyses (or transmitted and non-transmitted haplotype analyses) to investigate whether their effects are likely mediated through the fetal genome (i.e. direct effects of an individual’s genome on their own trait), the maternal genome (i.e. indirect effects of the mother’s genome mediated through e.g. the intrauterine environment on an individual’s trait) or some combination of both ^6,7^. Using this strategy, large-scale GWAS meta-analyses have identified over 280 variants robustly associated with a range of perinatal traits through both the fetal and/or maternal genomes ^1-6,8-10^.

Whilst this strategy of conducting separate fetal and maternal GWAS has been successful in terms of locus identification, it is not optimal statistically, because it does not utilize information shared across the individual GWAS. Since maternal and offspring genotypes are correlated, a GWAS of an individual’s own phenotype also provides information on indirect maternal genetic effects (i.e. in addition to information on direct fetal genetic effects). Likewise, GWASs of offspring phenotype also provide information on direct fetal genetic effects (i.e. in addition to indirect maternal genetic effects). Performing separate fetal and maternal GWAS therefore does not take full advantage of the information contained across both GWAS. This issue is most pronounced when the fetal and maternal GWAS are only partially overlapping (and hence contain independent information)- which has certainly been the case in the past for the GWAS meta-analysis of many phenotypes within the Early Growth Genetics (EGG) consortium including birth weight ^1-6^. However, it may not be clear how best to combine information from maternal and fetal GWAS, particularly when the degree of sample overlap/relatedness is unknown. In addition, when conducting two or more GWAS (e.g. GWAS of one’s own trait and then GWAS of the same trait in one’s offspring), investigators should ideally increase the statistical penalty due multiple testing-although this is not often done in practice and may not be easy to do optimally given unknown sample overlap and correlation between the traits. Data simulations and asymptotic power calculations we have performed previously using a computationally intensive structural equation model (i.e. that is not suitable for GWAS) have hinted at the gains in power that might be achieved by modelling and testing indirect and direct effects simultaneously in one analysis^7,11^.

In this manuscript we examine the performance of a computationally simple two degree of freedom test that uses GWAS summary results statistics from fetal and maternal GWAS. Our method uses LD score regression ^12^ to estimate an effective sample overlap across fetal and maternal GWAS, and then utilizes this information to estimate the sampling variance and covariance of the conditional direct fetal and indirect maternal genetic effect estimates. In addition, by essentially performing a single GWAS, the method avoids the thorny issue of how best to adjust for multiple testing across two correlated GWAS that contain an unknown proportion of overlapping/related individuals. We also compare the two degree of freedom test to a simple one degree of freedom test where signals from maternal and fetal GWAS are meta-analysed together conditional on the estimated sample overlap, as well as to the traditional strategy of separately analysing maternal and fetal GWAS. We investigate the power of the different approaches through a combination of analytical formulae and data simulation and provide users a web tool to conduct their own asymptotic power calculations. We apply the different methods to summary statistics from the most recent GWAS of birth weight involving the deCODE Study, EGG Consortium and UK Biobank^5^ and illustrate how combining the data dramatically increases the number of significantly associated loci for these traits relative to analysing the data separately or via the commonly used MTAG program^13^. We show how our methods can be extended to investigate the genetic aetiology of other phenotypes like educational attainment and IQ that are putatively influenced by indirect maternal and indirect paternal genetic effects as well as the direct effect from individuals’ own genomes^14-16^. Finally, we implement our methods in the DINGO (**<u>D</u>**irect and **<u>IN</u>**direct effects analysis of **<u>G</u>**enetic l**<u>O</u>**ci) software package, part of the online Complex Trait Genetics Virtual Laboratory (CTG-VL)(https://vl.genoma.io/)^17^, which allows users to perform these tests easily and computationally efficiently across the genome using summary results GWAS data.

## Method

### Formulation of the Two Degree of Freedom Test

Simply regressing own phenotype or offspring phenotype on own genotype will lead to inconsistent and biased estimates of association in the presence of indirect maternal and direct fetal genetic effects at a locus (see **Supplementary Note**). We and others have shown previously how consistent and unbiased estimates of indirect maternal and direct fetal genetic effects at individual variants can be obtained by combining summary statistic linear regression coefficient estimates of SNP-trait associations from maternal and fetal GWAS ^6,18^ i.e.:

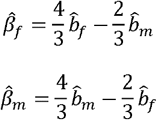

where 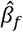 and 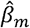 are estimates of the true population direct fetal (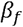) and indirect maternal genetic effects (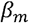), and 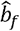 and 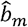 are linear regression coefficients from a GWAS of own phenotype on own genotype, and offspring phenotype on own (maternal) genotype respectively. The standard error of these terms can be estimated using the below formulae:

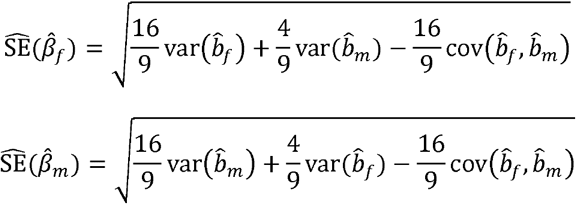

where “var” and “cov” refer to the sampling variance and covariance of the terms respectively ^6,18^. Note how the standard error of both terms is a function of the sampling covariance of 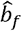 and 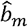. When GWAS of own and offspring traits are performed in separate unrelated families, this term can be assumed to be zero. However, this is rarely the case in practice. Wu et al. (2021)^18^ show how this term is a function of the sample overlap and the phenotypic correlation between overlapping mothers and children, and can be estimated using the intercept from bivariate LD score regression ^12^:

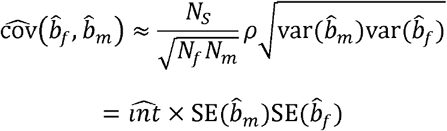

where *N*_*f*_ is the sample size of the GWAS of own genotype and own outcome (i.e. the “fetal GWAS”), *N*_*m*_ is the sample size of the GWAS of maternal genotype and offspring phenotype (i.e. the “maternal GWAS”), *N*_*s*_ is the effective number of overlapping individuals across both GWAS (e.g. in situations where all mothers report their own and their offspring’s birth weight the effective sample overlap would be 100%, whereas for genotyped mother-offspring pairs where only the offspring’s phenotype is measured, the effective sample overlap would be 50%, since offspring and maternal genotypes are correlated 0.5), and 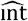 is the estimated bivariate LD score regression intercept. The parameter *ρ* refers to the correlation between own and offspring phenotype among the overlapping individuals.

In large samples, the hypothesis that the estimated maternal effect 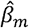 (or estimated fetal effect 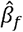) differs from zero can be evaluated for significance against the standard normal distribution i.e.:

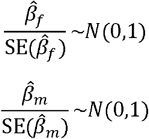

In this manuscript, we refer to these tests as “conditional one degree of freedom” tests. These conditional one degree of freedom tests have primary utility in locus characterization. That is, following the identification of the loci in GWAS, it is of interest to know whether the associated variant exerts its effect “directly” through the fetal genome, indirectly through the maternal genome, or some combination of both. However, we have shown that these conditional one degree of freedom tests often lack statistical power^7,11^ and typically do not produce stronger p-values than performing (potentially biased) unconditional tests of SNP-trait association in mothers or children separately. In other words, whilst conditional one degree of freedom tests are useful for locus characterization, in most situations they have limited utility for locus detection.

The joint sampling distribution of the estimated conditional maternal and fetal effects is bivariate normal in large samples i.e.:

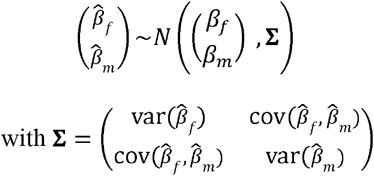

To increase the power of locus identification in GWAS of own and offspring phenotypes, we propose *T*_*2df*_, a two degree of freedom test that considers the joint distribution of the two effect estimates. It follows from elementary statistical theory that under the null hypothesis (*H*_*0*_) of no association between variant and maternal and fetal genotype (i.e. *β*_*f*_ = *β*_*m*_ = 0) the test statistic:

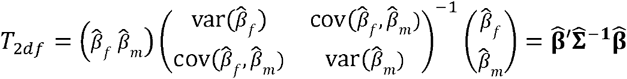

is asymptotically distributed as a chi-square statistic with two degrees of freedom (see the **Supplementary Note** for the derivation^19^). We verified using simulated data that *T*_*2df*_ conforms with our expectations under the null (**Supplementary Figure 1**).

This manuscript primarily focuses on perinatal traits like birth weight that are known to be influenced by variants in both the maternal and offspring genomes. However, we also derive an analogous three degree of freedom test in the **Supplementary Note** that could be useful for identifying loci influencing phenotypes like educational attainment that are likely to be a function of direct genetic, indirect maternal and indirect paternal effects.

### Formulation of One Degree of Freedom Meta-analytic Tests

We also investigated the power of a simple procedure for meta-analysing the results of (potentially overlapping) maternal and fetal GWAS. We illustrate this procedure in the case of fetal genetic effects noting that an analogous test for maternal genetic effects can also be derived in a complementary fashion. We first assume that maternal genetic effects are not present at the locus. Since maternal and fetal genotypes are correlated 0.5, we are able to obtain estimates of the fetal genetic effect from the maternal GWAS by simply doubling the estimated maternal effect regression coefficient (i.e. 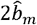), where 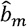 is the coefficient of the regression of offspring phenotype on maternal genotype. Likewise, the sampling variance of this estimate will be 4var (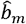). This estimate is then combined with estimates of the fetal genetic effect from the fetal GWAS by inverse variance weighted meta-analysis. The variance of this combined estimate is obtained by the usual inverse weighted variance formula plus an additional term due to twice the weighted covariance between the estimates:

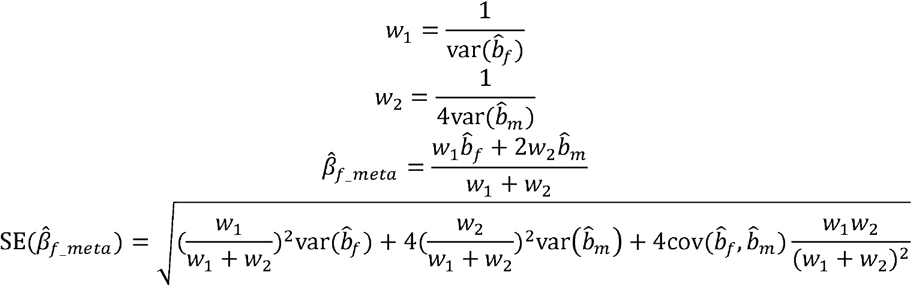

where 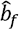 is the coefficient from the regression of own phenotype on own genotype and 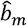 is the coefficient from the regression of offspring phenotype on maternal genotype, and 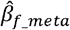 the inverse variance weighted estimate of the fetal effect across both these scans. The effective sample overlap between the scans is estimated using bivariate LD score regression and the covariance between the regression coefficients estimated from this quantity:

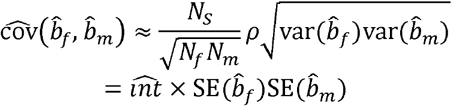

In large samples, under the null hypothesis of no association between SNP and trait (i.e. fetal or maternally mediated), the regression coefficients over their standard errors are distributed as standard normal:

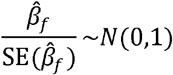

We refer to this test/procedure as the one degree of freedom meta-analytic strategy. We note that when both maternal and fetal effects are present at a locus, simply meta-analysing the data as described above will result in biased estimates of maternal and fetal genetic effects. However, since our interest is on locus discovery (i.e. regardless of whether a genetic effect is mediated by the fetal or maternal genome), we argue that this bias can be ignored, as the type I error for both tests should be well calibrated under the null (i.e. when neither maternal nor fetal effects contribute to trait variation at the locus). Indeed, if desired, unbiased estimates of maternal and fetal genetic effects can be obtained post-hoc using the two degree of freedom procedure described in the previous section.

With a couple of notable exceptions (e.g. the Norwegian MOBA cohort), the majority of the world’s birth cohorts contain only limited genotype information on the children’s fathers. Nevertheless, in some situations it may be useful to include information from a GWAS of offspring phenotype regressed on father’s genotype. This could be useful when paternal effects are suspected to contribute to offspring trait variation, or (in their absence) when fathers provide information on their offspring in the absence of offspring genotype information (i.e. in which case the fathers are providing information indirectly on offspring mediated effects). In the **Supplementary Note** we derive analogous formulae for meta-analyses when paternal, maternal, and fetal GWAS are available.

### Power

We performed extensive data simulations to investigate the power of the three locus detection strategies described in this manuscript i.e. (i) the usual strategy of separately analysing maternal and fetal GWAS with one degree of freedom tests; ii) two degree of freedom *T*_*2df*_ tests applied to the maternal and fetal GWAS concurrently; and (iii) meta-analysing maternal and fetal GWAS to obtain estimates of the maternal and fetal effect using one degree of freedom tests. We simulated bivariate normally distributed quantitative traits of unit variance for own and offspring phenotype that were influenced by a single additive locus (N = 1000 simulations). We varied locus effect size (three conditions: (i) *β*_*f*_ = 0.1 and *β*_*m*_= 0.1; (ii) *β*_*f*_ = 0 and *β*_*m*_ = 0.1; (iii) *β*_*f*_ = -0.1 and *β*_*m*_ = 0.1), residual correlation between maternal and fetal phenotype (three conditions: (i) *ρ* = 0; (ii) *ρ* = 0.25; (iii) *ρ* = 0.5), sample size of each GWAS (19 conditions: N = 100 to 900, increment by 100, and then N = 1,000 to 10,000, increment by 1000), relative sample size (two conditions: (i) equal sized maternal and fetal GWAS; (ii) maternal GWAS half the size of fetal GWAS) and extent of sample overlap (three conditions: (i) zero overlap; (ii) 50% of individuals in the maternal GWAS in the fetal GWAS; (iii) all of the mothers in the maternal GWAS in the fetal GWAS).

Since the locus detection strategies involving one degree of freedom tests entail performing double the number of statistical tests relative to the two degree of freedom test (i.e. performing genome-wide tests for a fetal effect, and genome-wide tests for a maternal effect), we evaluated power for the one degree of freedom tests against a genome-wide Bonferroni corrected alpha level of α/2 = 6.6 × 10^−9^/2 = 3.3 × 10^−9^ (for a justification of why a Bonferroni correction is reasonable, see the **Supplementary Note**). However, since our focus is on locus discovery, and so technically we are agnostic as to whether evidence for association comes from the maternal or fetal meta-analysis, we evaluated power according to whether *either* the maternal *or* the fetal test was significant for a given replicate. Note that for our data simulations we adopted a slightly more stringent significance level than the typical α = 5x10^−8^ threshold employed in most GWAS because of the increased number of low frequency variants contained within modern imputation panels (see Morris et al (2019) for further justification of this threshold)^20^.

In the **Supplementary Note** we derive analytical expressions for the non-centrality parameter (and hence power) of all the statistical tests described in this manuscript and confirm that the asymptotic power implied by the expressions matches the results of our simulations closely. We encode several of our asymptotic formulae into an online web calculator that allows users to approximate power in their own GWAS (https://evansgroup.di.uq.edu.au/DINGO).

### Empirical application

We applied our strategies to birth weight GWAS summary results statistics from the most recent GWAS of birth weight^6^ which contains meta-analysed data from deCODE, the EGG Consortium and the UK Biobank. The GWAS of own birth weight included 423,683 individuals of European ancestry and the GWAS of offspring birth weight included 270,002 women of European ancestry as defined by the authors of the study (https://www.decode.com/summarydata/). Given that the standard errors were not provided in the GWAS summary statistics, we derived them using p-values and beta effect estimates and the qnorm function in R, i.e. standard error = abs(beta / qnorm(p-value / 2)). Among 33,409,268 single nucleotide polymorphisms (SNPs) from the GWAS of own birth weight and 33,453,671 SNPs from the GWAS of offspring birth weight, 1,276,496 and 1,220,850 SNPs were removed, respectively, due to p-value = 1 or beta = 0 because in either of these conditions standard errors cannot be derived. For the remaining SNPs, 31,033,794 were available in both GWAS and were included in the two degree of freedom *T*_*2df*_ test and in the meta-analyses of the estimated fetal and maternal effects. We performed clumping analyses for SNPs within a 500-kb window size using r^2^ of 0.05, and α = 6.6 × 10^−9^, and the 1000 Genome Phase 3 EUR reference panel using the online platform FUMA^21^. We compared genome-wide significant loci from the two degree of freedom test against those from the published GWAS that used a one degree of freedom test (evaluated at α = 3.3 × 10^−9^) and with loci identified in the meta-analyses of maternal/fetal effects (evaluated at α = 3.3 × 10^−9^). For genome-wide significant SNPs, we then applied conditional one degree of freedom tests (evaluated at α = 0.05) to test for whether effects were likely to be mediated through the maternal and/or fetal genomes. Finally, for lead variants at loci not reported as genome-wide significant in the most recent deCODE meta-analysis of birth weight^5^, we searched the GWAS catalogue and variants in strong linkage disequilibrium (r^2^ > 0.9) for reported trait associations using the FUMA software package^21^.

### Comparison with other multivariate software

*T*_*2df*_ differs from other commonly used multi-trait association analyses that do not precisely model the correlation between the parents and their offspring. We therefore compared the performance of *T*_*2df*_ against MTAG (evaluated at α = 3.3 × 10^−9^), a commonly used tool for boosting statistical power to detect loci underlying correlated traits using summary results statistics ^13^. We applied MTAG to the same summary results statistics of fetal and maternal birth weight as described above and compared the results against those from *T*_*2df*_.

## Results

### Power analyses

**Figure 1** and **Supplementary Figures 2 – 6** compare the power to detect association (as assessed by simulation) across the three analytic strategies examined in this manuscript. When there was minimal sample overlap between maternal and fetal GWAS, then the usual strategy of analysing maternal and fetal GWAS separately was typically less powerful than strategies that combined information from both GWAS (**Figure 1** bottom panel). In general, in these situations, the meta-analysis strategy was often slightly more powerful than the two degree of freedom test in detecting loci that exhibited one type of effect only (**Figure 1** bottom middle panel) and when detecting loci with concordant maternal and fetal effects (**Figure 1** bottom left panel).

**Figure 1.**
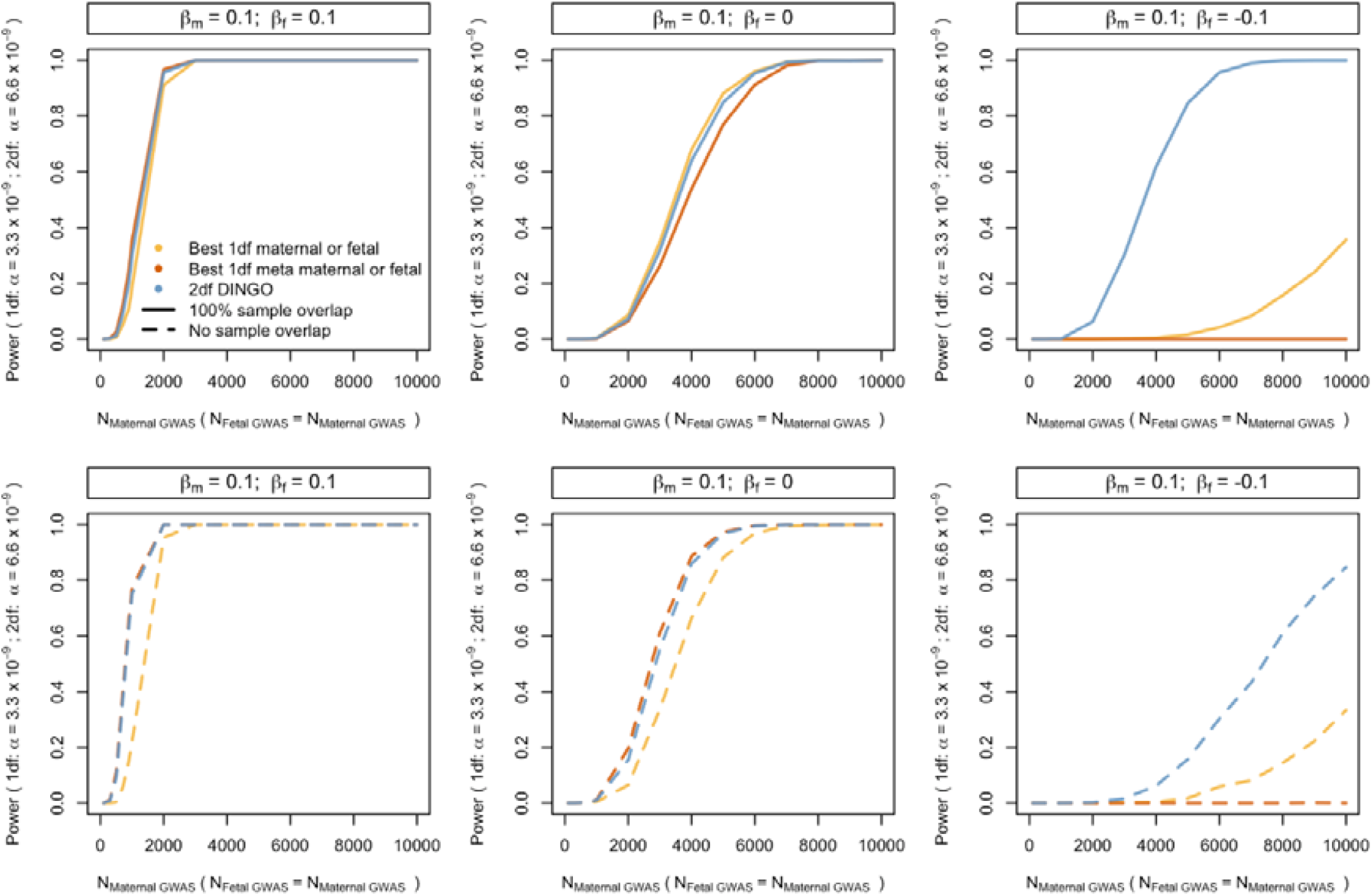
Power to detect association (as evaluated using simulation) using a traditional strategy of analysing separate maternal and fetal GWAS, a strategy involving one degree of freedom meta-analyses, and a strategy of performing two degree of freedom *T*_2df_ tests across the genome. β_m_ and β_f_ refer to maternal and fetal genetic effects on a standardized trait. In the top row, results are shown for replicates where there is complete sample overlap. Data were simulated assuming a high residual correlation between maternal and offspring phenotypes (*ρ* = 0.5). In the bottom row, results are shown for replicates where there is no sample overlap. For the traditional strategy of running separate maternal and fetal GWAS and the one degree of freedom meta-analysis strategy, we set the alpha value to α = 3.3 × 10^−9^, i.e. half the α = 6.6 × 10^−9^ type I error rate of the two degree of freedom *T*_*2df*_ test in order to take into account that we are performing twice the number of statistical tests in the former situations. In the case of the traditional strategy of running separate maternal and fetal GWAS and the one degree of freedom meta-analysis strategy, we evaluated power with respect to whether *either* test met the criterion for genome-wide significance for each replicate.

In contrast, at the other extreme, when there was complete sample overlap, then the optimal strategy depended upon whether the locus exhibited both type of effects concurrently and the residual correlation between variables. For example, **Figure 1** (middle panel) shows an instance of where the usual strategy of analysing the maternal and fetal GWAS separately was most powerful-in this case where loci exhibited only indirect maternal (or only direct fetal) genetic effects and in which the residual correlation was very high. In contrast, when loci exhibited concordant maternal and fetal effects of similar magnitude then both the two degree of freedom test and the meta-analysis strategy often gave superior power compared to separately analysing the scans (**Figure 1** top left panel).

Interestingly, in the case of loci that exhibited discordant maternal and fetal effects (of similar magnitude), the two degree of freedom *T*_*2df*_ test provided increased power to detect association relative to the other strategies, particularly in overlapping samples (**Figure 1** top right panel). Notably, the simple meta-analysis strategy did not perform well in these situations (see **Supplementary Table 2** for further details).

In the **Supplementary Note** we derive asymptotic formulae for the unconditional GWAS tests, the meta-analytic strategy, and the two degree of freedom tests of association. We confirm that our formulae for asymptotic power of the two degree of freedom test derived in the **Supplementary Note** gave similar results to those obtained using simulated data (results not shown) and implement our calculations in a web calculator (https://evansgroup.di.uq.edu.au/DINGO) that investigators can use to investigate power in their own studies.

### Empirical analyses of birth weight using the two degree of freedom test

The GWAS of birth weight using the two degree of freedom *T*_*2df*_ test identified 332 independent SNP associations at 213 loci with a p-value < 6.6 × 10^−9^ (**Figures 2 and 3, Supplementary Table 3**). This included 34 independent SNPs at 32 loci that did not reach genome-wide significance in either the separate GWAS of birth weight or offspring birth weight (**Supplementary Table 3**). These results are consistent with the preceding power calculations which suggest that performing a two degree of freedom test can be substantially more powerful than performing separate maternal and fetal GWAS when there is only partial overlap between the GWAS. Conditional analyses suggested that among 34 genome-wide significant SNPs at the 32 loci, 10 acted primarily through the fetal genome, 12 acted primarily through the maternal genome, and 12 acted through both (p-value < 0.05) (**Figure 4**). Interestingly, of the 12 SNPs that showed evidence for both maternal and fetal effects in the conditional analyses, all 12 SNPs exhibited effects in the same direction (**Figure 4**). These results were at first surprising, given that the power calculations from the previous section suggested that whilst the two degree of freedom test should increase the power to detect loci that show concordant maternal and fetal effects when there is partial sample overlap, the largest gains in power should involve the detection of SNPs that have discordant maternal and fetal effects of similar magnitude. Indeed only 22 of 332 genome-wide significant SNPs identified using the two degree of freedom test exhibited conditional maternal (p-value < 0.05) and fetal effects (p-value < 0.05) in opposite directions- and all these SNPs were also identified in the separate GWAS analyses of own or offspring birth weight. **Figure 5** shows the likely reason for this observation-most birth weight associated SNPs that exhibited discordant maternal and fetal genetic effects have estimated maternal and fetal effect sizes of quite different magnitude. Indeed, although there are some SNPs that show sizeable (discordant) maternal and fetal effects, these effects were so large that they were also picked up by one degree of freedom tests in the separate GWAS of birth weight and offspring birth weight. In contrast, there were many SNPs that showed concordant maternal and fetal effects of similar magnitude and in many of these cases the two degree of freedom test was more powerful than separate GWAS (**Figure 5**).

**Figure 2.**
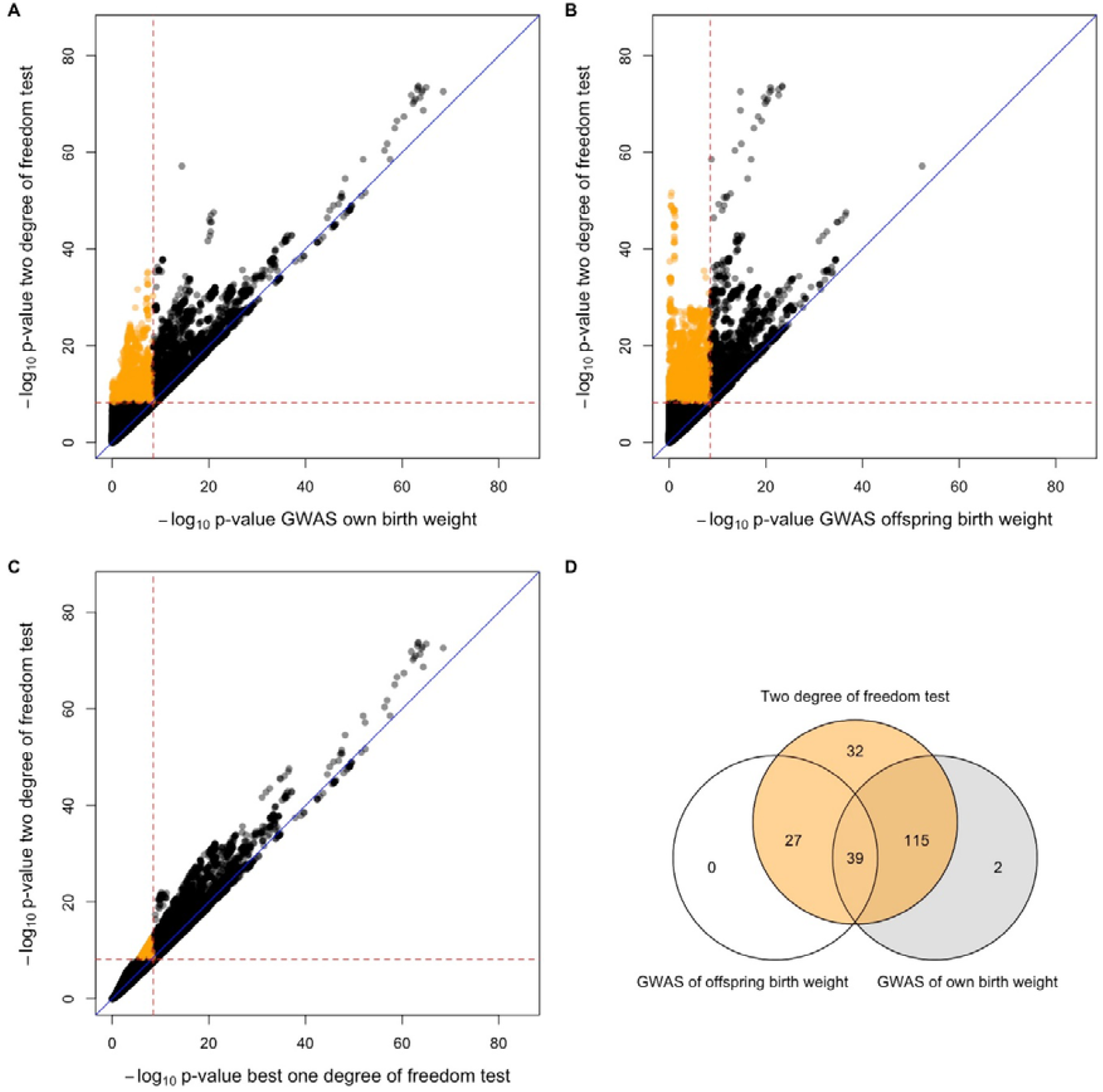
P-value scatter plots and Venn diagram comparing results from separate GWAS of own and offspring birth weight versus the combined two degree of freedom test across 31,033,794 SNPs from the deCODE study of birth weight. The -log_10_p-value from the two degree of freedom test (y-axis) was compared against the -log_10_p-value of SNPs from (A) the separate GWAS of own birth weight, (B) the separate GWAS of offspring birth weight, and (C) the stronger p-value from among A and B (x-axis). Red dashed lines denote genome-wide significant thresholds of α = 3.3 × 10^−9^ for the separate GWAS of own birth weight and offspring birth weight which involve one degree of freedom tests, and α = 6.6 × 10^−9^ for the two degree of freedom test. The blue diagonal lines indicate x = y. Orange circles are SNPs that are only genome-wide significant in the two degree of freedom test. Panel (D) illustrates the degree of overlap across genome-wide significant loci identified from the two strategies.

**Figure 3.**
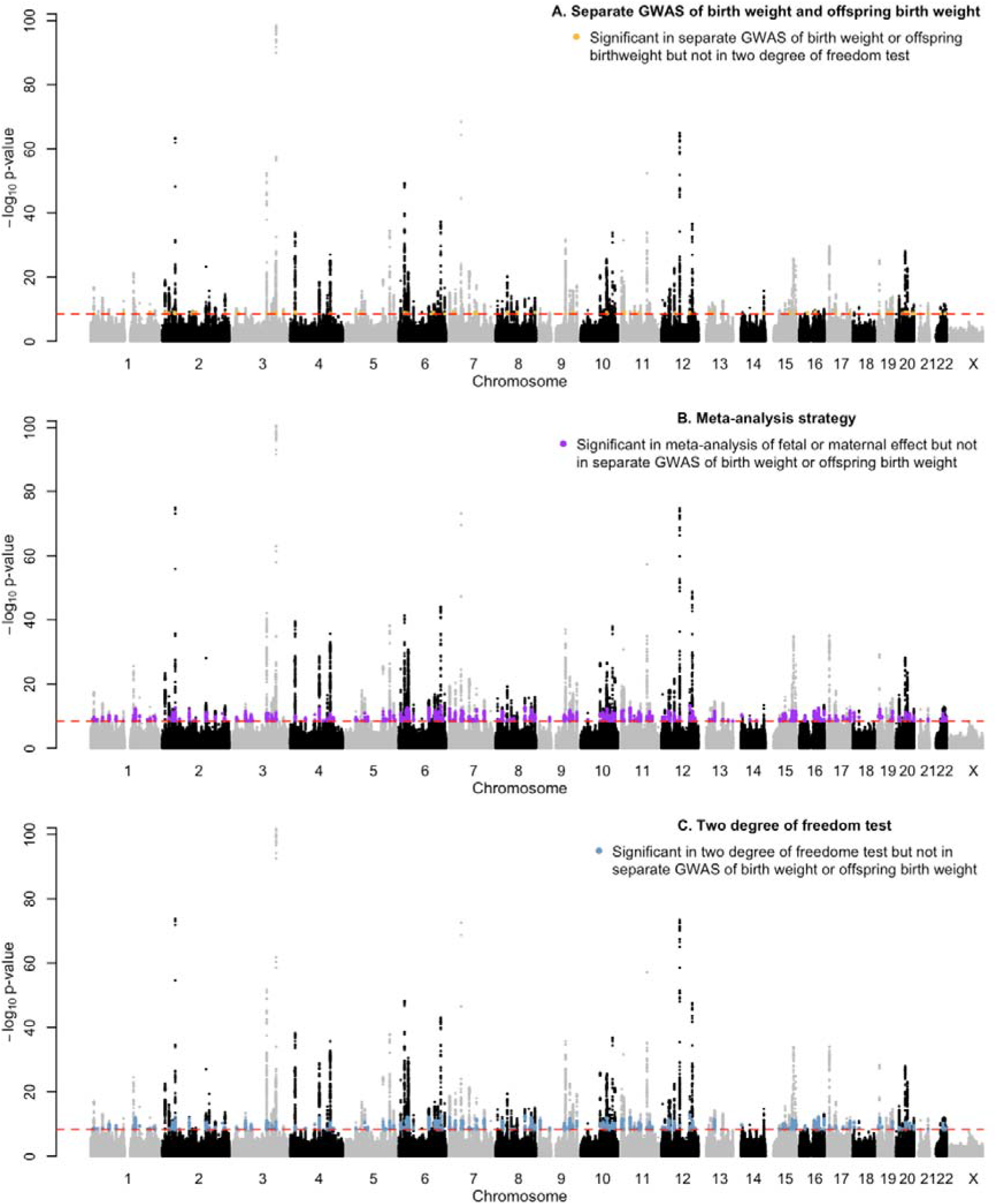
Manhattan plots showing results of using the typical strategy of conducting separate GWAS of birth weight and offspring birth weight (top panel-best p-value at each locus shown), a simple meta-analytic strategy (middle panel-best p-value at each locus shown), and the two degree of freedom test (bottom panel). The -log_10_ p-value for the SNP association is plotted on the y-axis. Red dashed lines indicate genome-wide significant thresholds of p-value = 3.3 × 10^−9^ for one degree of freedom tests, and p-value = 6.6 × 10^−9^ for the two degree of freedom tests. Orange points represents SNPs that are genome-wide significant in the separate GWAS of birth weight/offspring birth weight, but not in the two degree of freedom test. Purple points represent SNPs that are genome-wide significant in the meta-analysis of fetal or maternal effects, but not in the separate GWAS of birth weight/offspring birth weight. Blue points represent SNPs that are genome-wide significant in the two degree of freedom test, but not in the separate GWAS of birth weight/offspring birth weight.

**Figure 4.**
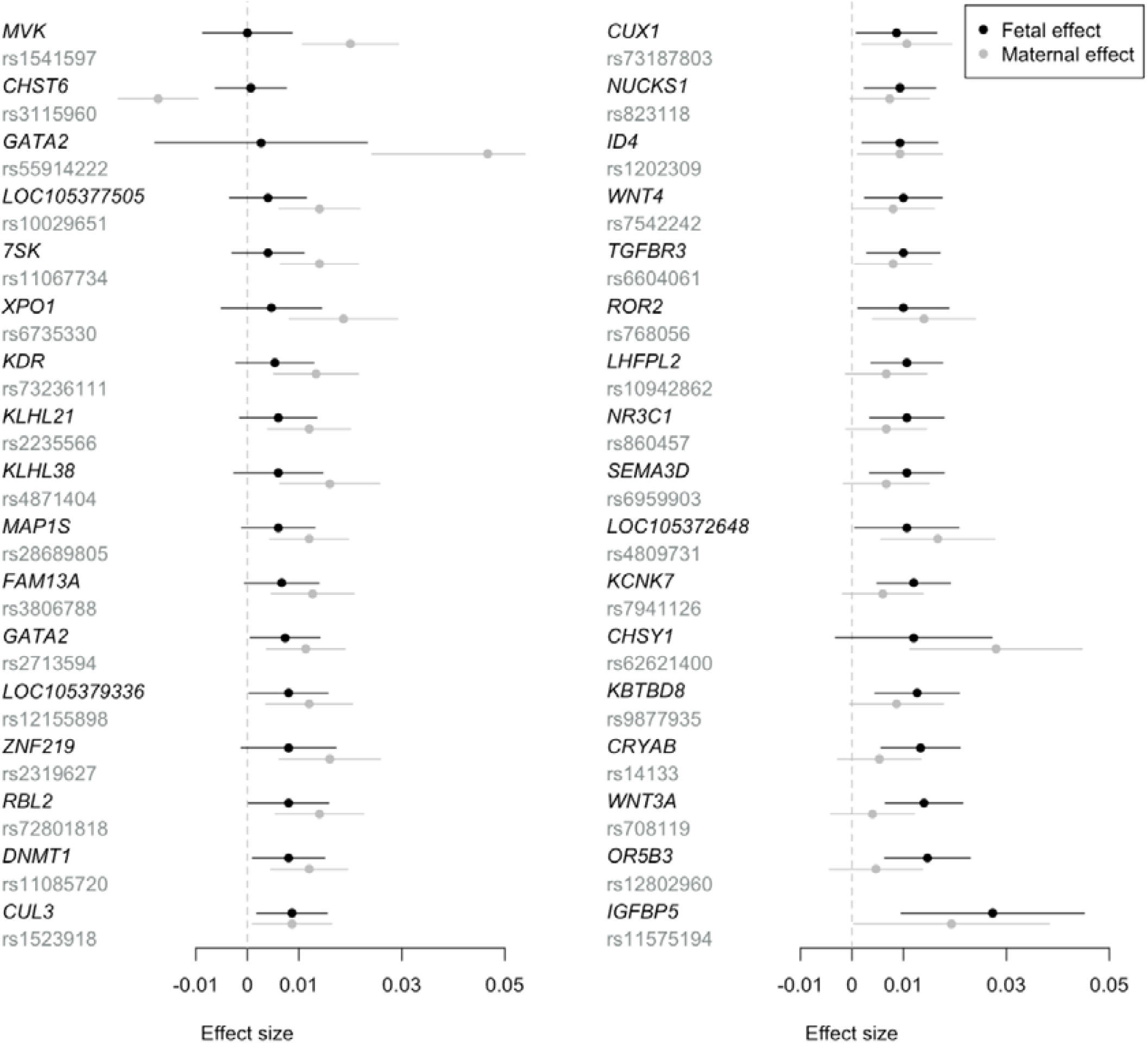
Estimated conditional fetal and maternal effects and 95% confidence intervals for the 34 independent genome-wide significant SNPs identified in the two degree of freedom analysis (p-value < 6.6 × 10^−9^) that did not meet genome-wide significance in the separate GWAS of birth weight and offspring birth weight (p-value < 3.3 × 10^−9^). Effects are oriented to the allele that is associated with an increased fetal effect on birth weight. The SNPs are labelled according to the physically closest gene. The upper 95% confidence interval for rs55914222 has been truncated.

**Figure 5.**
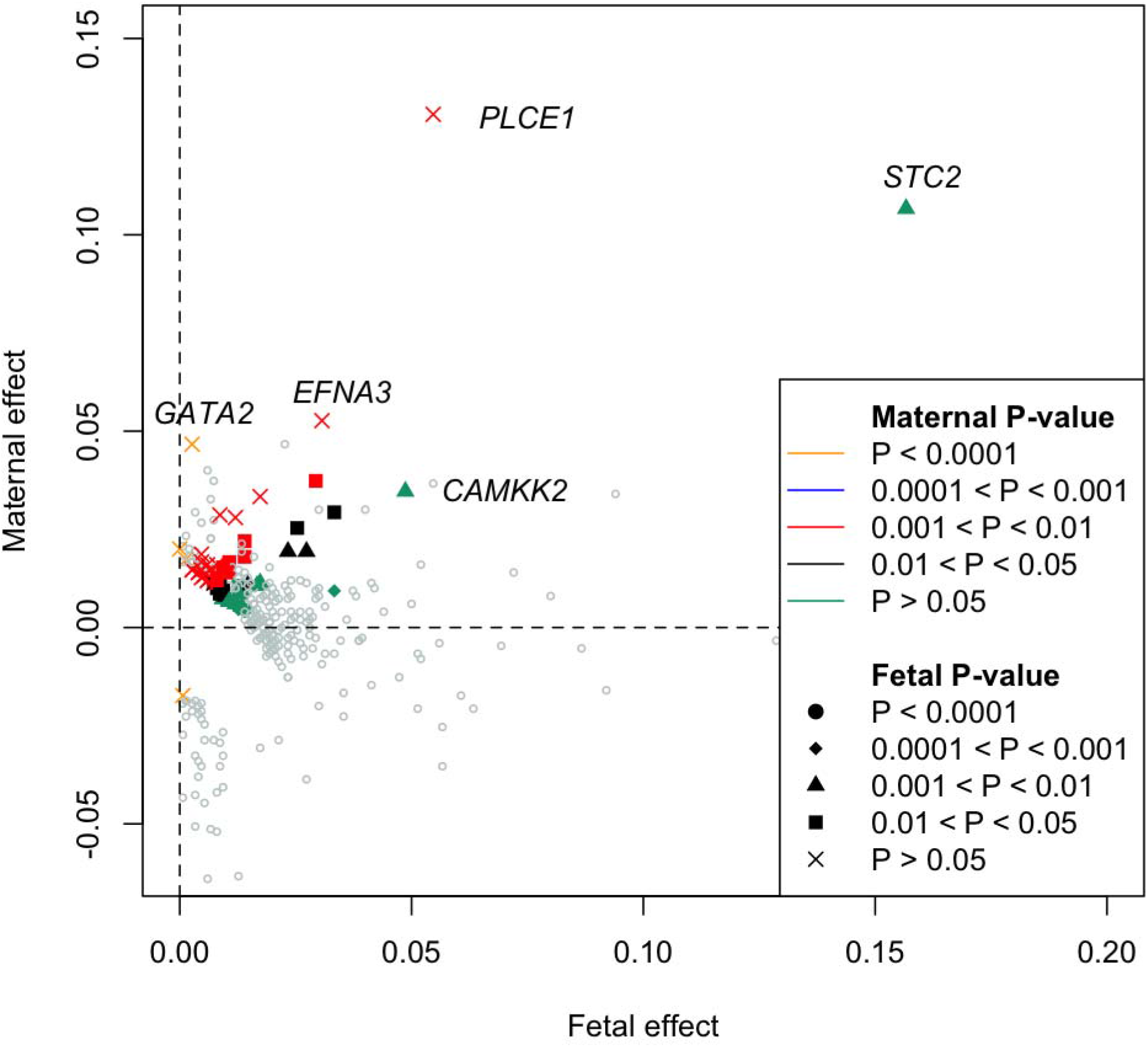
Estimates of conditional fetal and maternal effects for 332 genome-wide significant SNPs identified in the two degree of freedom GWAS. 76 SNPs* that reached genome-wide significance in the two degree of freedom test (p-value < 6.6 × 10^−9^), but not in either the GWAS of birth weight or offspring birth weight (p-value < 6.6 × 10^−9^) are displayed in colour, whilst the other 256 SNPs are shown in grey. Effect sizes and p-values represent conditional one degree of freedom fetal and maternal tests calculated from the GWAS of birth weight and offspring birth weight using DINGO. The colour of each point represents the p-value for the conditional maternal effect and the shape of each point the p-value for the conditional fetal effect. SNPs with large conditional effects are labelled with the name of the closest gene. Note in particular the paucity of SNPs that show discordant maternal and fetal effects of similar magnitude. *It is possible that other SNPs at the same loci as these SNPs reached genome-wide significance in the GWAS of birth weight or offspring birth weight

There were five (one) SNPs that reached genome-wide significance in the GWAS of own birth weight (offspring birth weight) (p-value < 3.3 × 10^−9^) but did not reach significance in the two degree of freedom GWAS (p-value < 6.6 × 10^−9^), however, only two of these (both from the GWAS of own birth weight) involved loci not identified by the two degree of freedom test. All these SNPs exhibited either predominantly fetal or maternal effects only (**Supplementary Table 4**).

Finally, the two degree of freedom test outperformed the one degree of freedom tests implemented in the popular multivariate GWAS package MTAG (**Supplementary Figure 7**), which only identified 23 novel independent significant SNP associations with p-value < 3.3 × 10^−9^, all of which were identified in both the 2 degree of freedom *T*_*2df*_ test and the one degree of freedom meta-analyses described in the next section (**Supplementary Table 5**). In conditional analyses, all 23 of these SNPs had estimated maternal and fetal effects in the same direction.

### Empirical analyses of birth weight using one degree of freedom meta-analyses

The one degree of freedom meta-analytic strategy identified 369 independent SNPs at 226 loci that were genome-wide significant (p-value < 3.3 × 10^−9^) in either the meta-analysis of fetal effects, or the meta-analysis of maternal effects – an even greater number of loci than were identified using the two degree of freedom strategy (**Supplementary Table 6**). These included 21 independent genome-wide significant SNPs at 21 loci that were not identified in the two degree of freedom *T*_*2df*_ tests (**Figure 6, Supplementary Table 6**), although all only narrowly missed out on significance in the two degree of freedom *T*_*2df*_ test, having a p-value smaller than 5 × 10^−8^. Interestingly, eight independent SNPs (at eight loci) were significant in the *T*_*2df*_ test but not in the meta-analysis of fetal effects, or the meta-analysis of maternal effects (**Figure 6, Supplementary Table 3, Supplementary Table 6**). Four of these SNPs exhibited significant (p-value <0.05) discordant maternal and fetal effects in the conditional analyses– most notably rs7034200 at *GLIS3*, a known type 2 diabetes locus, that was highly significant in the two degree of freedom test (p-value_2df_ = 1.15 × 10^−12^), but only exhibited relatively modest non-significant p-values in the maternal and fetal meta-analyses (minimum p_meta_ = 7.39 × 10^−5^), and likewise rs560887 at *G6PC2*, a known locus for fasting glucose which also exhibited radically different p-values across the two tests (p-value_2df_ = 4.37 × 10^−20^, minimum p_meta_ = 6.25 × 10^−7^).

**Figure 6.**
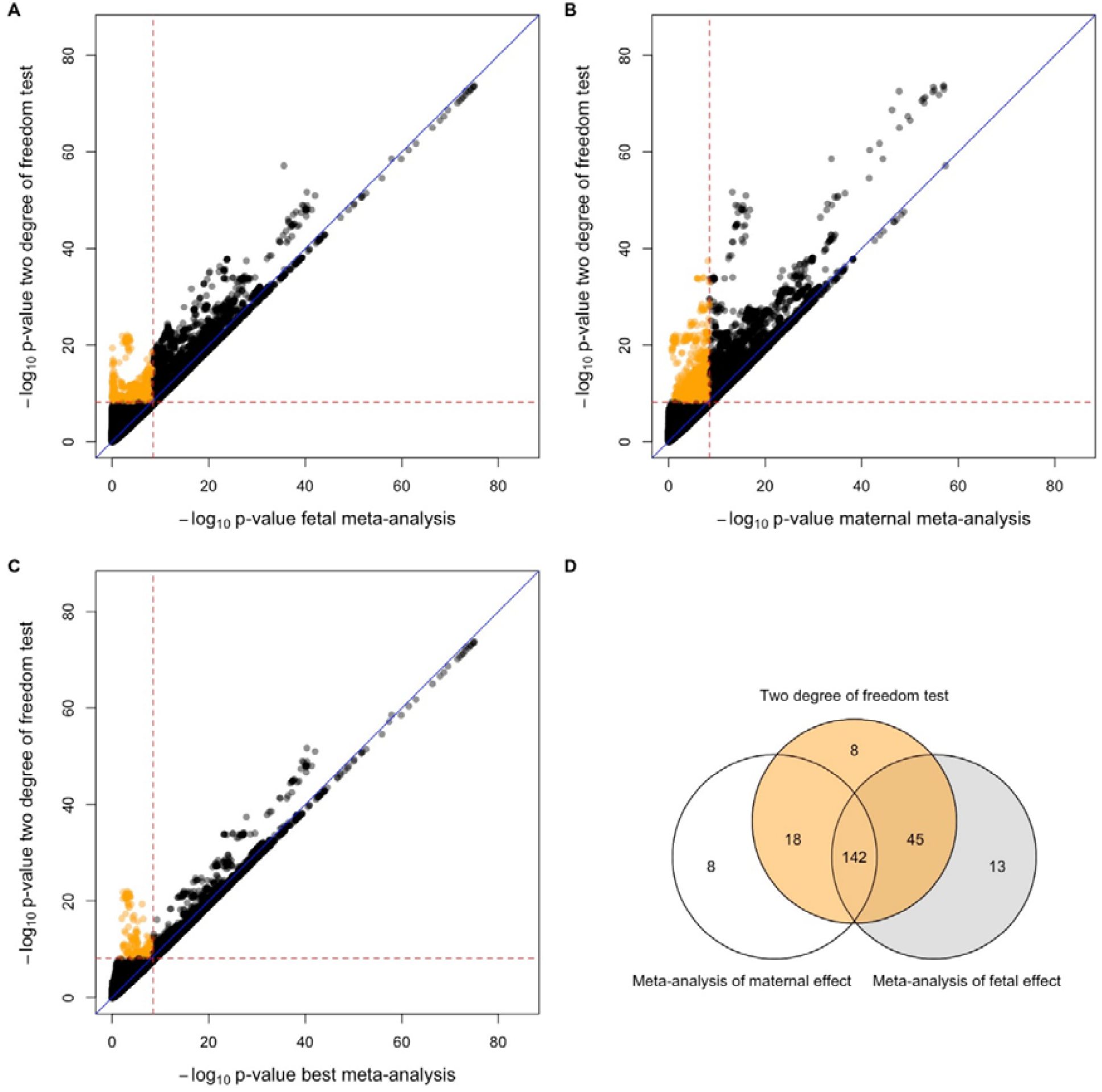
Quantile-Quantile plots and Venn diagram comparing one degree of freedom meta-analysis tests versus the combined two degree of freedom DINGO test for birth weight across 31,033,794 SNPs. The -log_10_p-value of the two degree of freedom DINGO test calculated using the GWAS of fetal birth weight and the GWAS of maternal birth weight was compared against the -log_10_p-value of SNPs from (A) the GWAS meta-analysis of fetal birth weight, (B) the GWAS meta-analysis of maternal birth weight, and (C) the stronger p-value from among A and B. Red dashed lines denote to genome-wide significant thresholds of α = 3.3 × 10^−9^ for the one degree of freedom maternal and fetal tests and α = 6.6 × 10^−9^ for the two degree of freedom DINGO test. Blue diagonal lines indicate x=y. Orange circles are SNPs that are only genome-wide significant in the two degree of freedom test. There are a considerable number of SNPs that reach genome-wide significance in the GWAS meta-analysis but in the not the two degree of freedom test. These SNPs are all clustered in a very small region of Figures 6a, 6b and 6c in the bottom right quadrant adjacent to the thresholds for genome-wide significance. (D) illustrates the overlap between genome-wide significant loci identified from one and two degree of freedom tests.

We compared the results of our one degree of freedom meta-analyses against the list of variants claimed as genome-wide significant in the most recent GWAS analysis of birth weight^5^. Our analyses identified an additional 68 SNPs at 62 novel loci compared to the deCODE paper (**Table 1**). These included many SNPs previously associated with anthropometric traits (height, body mass index etc), blood pressure, bone mineral density and type 2 diabetes at genome-wide significant levels (**Supplementary Table 7**). These are all phenotypes that are known to be causally or pleiotropically associated with birth weight, increasing our confidence that our new findings represent genuine associations. Finally, we also found several new associations between SNPs and birth weight that could implicate new biological pathways in the aetiology of the trait. These include an interesting association between rs11085720, an intronic variant in the DNA methylase transferase 1 (*DNMT1*) gene, and birth weight.

**Table 1.**
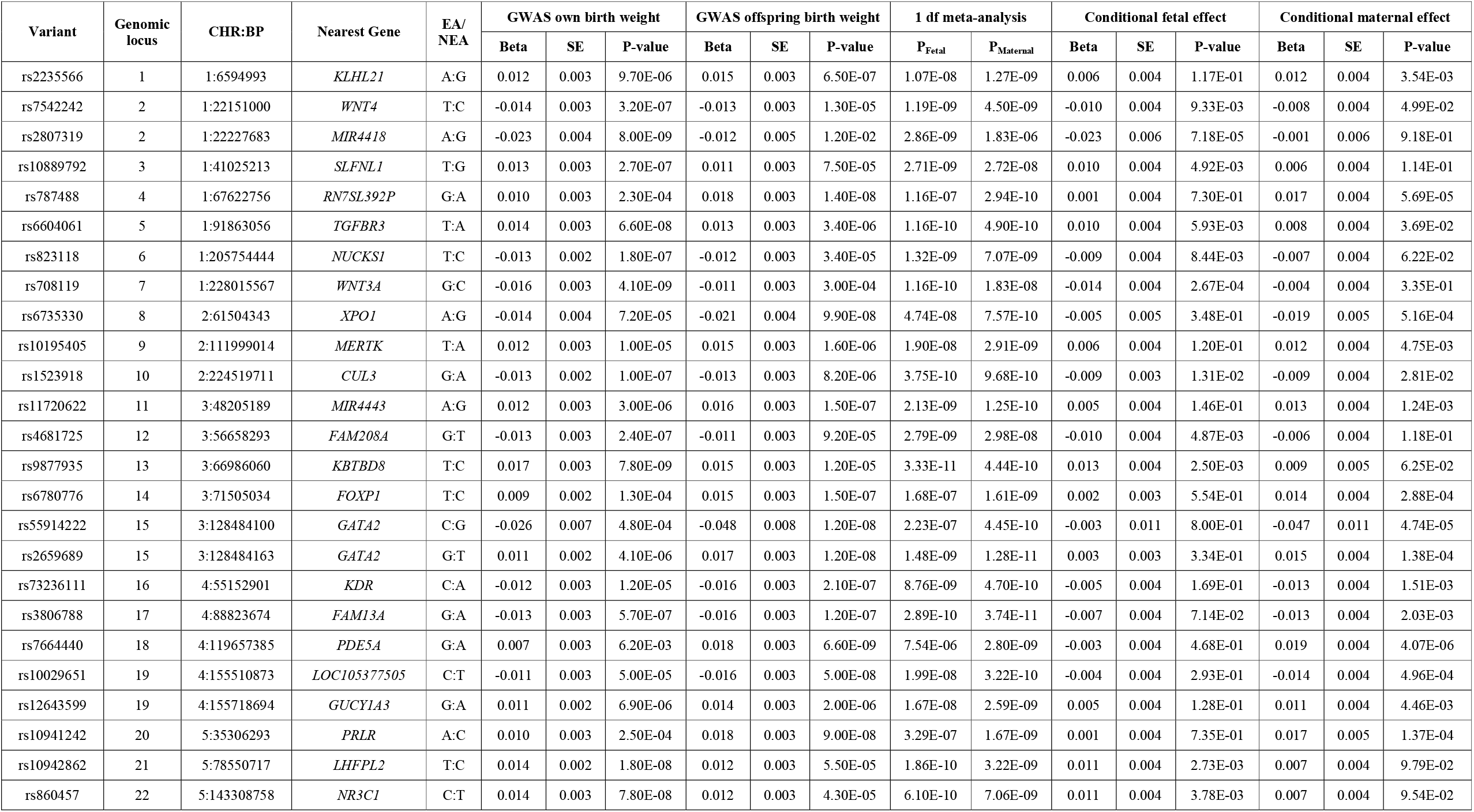
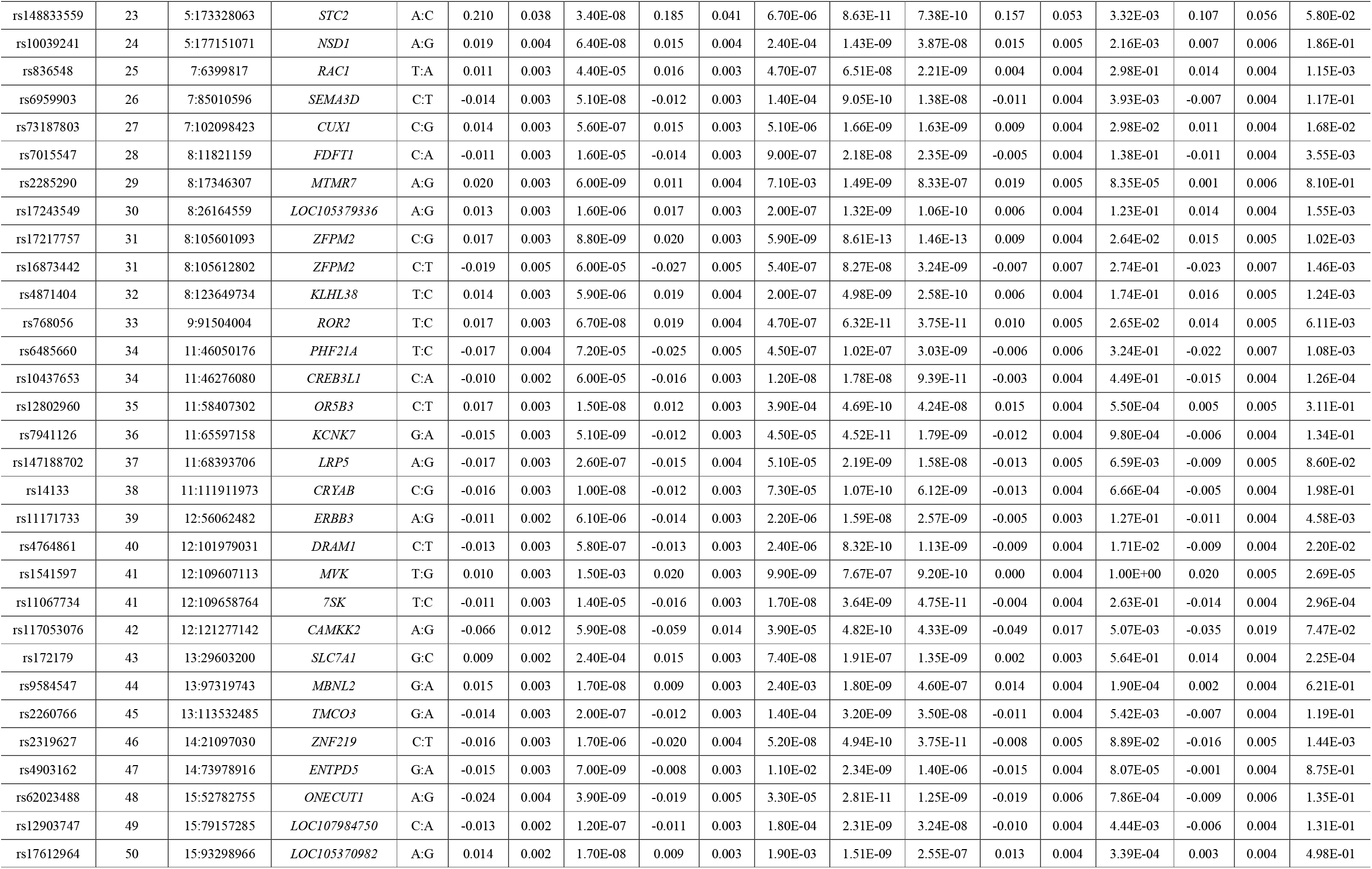
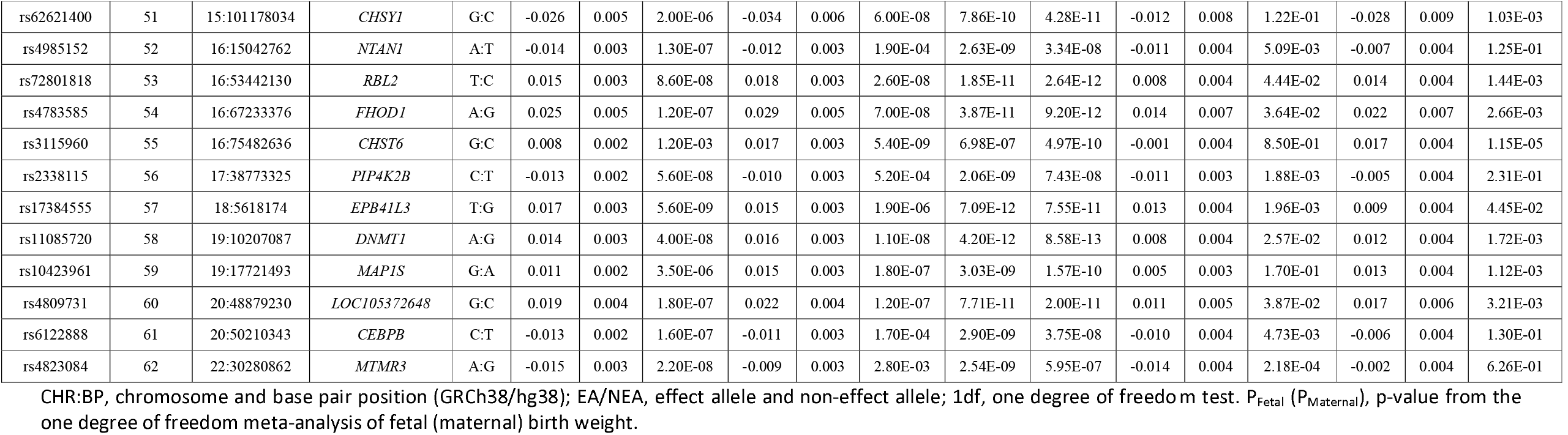
Novel genetic variants for birth weight identified in the one degree of freedom meta-analyses. We identified 68 novel independent genome-wide significant associations (p-value < 3.3x10^−9^) at 62 genetic loci that were at least 500kb apart from those previously reported in the latest deCODE analysis of birth weight ^5^. Genome-wide significance in the deCODE study was determined using class-based significance thresholds i.e. p-value < 2.49 × 10^−7^ for high impact variants; p-value < 4.97 × 10^−8^ for moderate impact variants; p-value < 4.52 × 10^−9^ for low-impact variants, p-value < 2.26 × 10^−9^ for other low-impact variants in DNase I hypersensitivity sites and p-value < 7.53 ×10^−10^ for all other variants.

## Discussion

In this manuscript we compared the power of three strategies to detect loci in maternal and fetal GWAS of the same trait: (i) the traditional strategy of analysing maternal and fetal GWAS separately; (ii) a two degree of freedom test which combines information from maternal and fetal GWAS; and (iii) a one degree of freedom test where signals from maternal and fetal GWAS are meta-analysed together conditional on the estimated sample overlap. We found that which strategy performed best depended on the degree of sample overlap between maternal and fetal GWAS, the correlation between own and offspring phenotype, whether individual trait loci were simultaneously influenced by maternal and fetal effects, and if so, whether the effects were directionally concordant.

Our simulations showed that in general, when maternal and fetal GWAS contained only partially overlapping samples (e.g. as is the case for most published GWAS meta-analyses of perinatal traits to date), considerable gains in power could be achieved using the meta-analytic strategy or using the two degree of freedom test compared to the traditional approach of analysing the scans separately. This makes sense intuitively since the traditional strategy of performing separate GWAS does not make use of the information contained across the scans. In these situations, for loci exhibiting concordant maternal and fetal effects (of similar magnitude) the two degree of freedom and the meta-analytic strategy typically performed similarly, whereas for loci involving only maternal or only fetal effects, a simple meta-analysis was often most powerful. Our results imply that many existing GWAS for perinatal traits could benefit from reanalysis using a two degree of freedom test or meta-analytic strategy (a point we demonstrate in this manuscript with the empirical analysis of birth weight).

In contrast, when maternal and fetal GWAS involved completely overlapping samples (as is the case for cohorts where all individuals report their own and their offspring’s phenotype), then which strategy was best depended on whether individual trait loci were simultaneously influenced by maternal and fetal effects, whether the effects were directionally concordant, and the background correlation between own and offspring phenotype. For loci that only exhibited effects through the fetal or maternal genome, then the traditional strategy of analysing maternal and fetal GWAS separately performed best. For loci that exhibited concordant maternal and fetal effects, the meta-analytic strategy was best (with the two degree of freedom test a close second).

Interestingly, our simulations also showed that when a single locus exhibited maternal and fetal effects that operated in different directions, a two degree of freedom test that modelled these effects often had vastly improved power to detect association compared to simple one degree of freedom tests (regardless of the degree of sample overlap). This result is similar to what we have observed previously in tests of genetic association involving individual level genotypes using a structural equation modelling framework^7,11^. Our results make sense intuitively as the two degree of freedom test expends an extra degree of freedom productively modelling both maternal and fetal effects, whereas the meta-analytic strategy combines effects in such a way that discordant effects may cancel out. Loci that exhibit maternal and fetal genetic effects in opposite directions are not uncommon in GWAS of perinatal traits related to growth ^6,7^. The implication of our power analyses is that a proportion of these loci may be missed if only one degree of freedom tests are performed across the genome (i.e. either by meta-analysis or separate maternal and fetal GWAS).

In empirical analyses of birth weight, and consistent with our simulations, both the two degree of freedom test and the meta-analytic strategy identified greater numbers of loci meeting genome-wide significance than analysing maternal and fetal GWAS separately. Both the meta-analytic strategy and the two degree of freedom test also identified more genome-wide significant loci for birth weight than MTAG ^13^, a popular software package that is commonly used in the multivariate analysis of GWAS summary results statistics. This makes sense since the two degree of freedom test (and the meta-analytic strategy) are derived by specifically modelling the correlation between maternal and fetal genotypes whereas other multivariate tests e.g. MTAG^13^ use models that are agnostic to the mother-offspring relationship and do not take advantage of this known source of information.

So which type of analysis is optimal in the case of birth weight and (potentially) other perinatal phenotypes, particularly in situations where there is only partial overlap between maternal and fetal GWAS? The meta-analytic strategy yielded considerably greater numbers of loci meeting genome-wide significance than the two degree of freedom strategy (i.e. 226 vs 213 loci), suggesting that even for a trait like birth weight, a simple meta-analytic strategy may be optimal when maternal and fetal GWAs are only partially overlapping. However, whilst all loci that met genome-wide significance using the simple meta-analytic strategy were also significant or almost significant using DINGO (i.e. all loci would be flagged by researchers as significant or suggestive), many genome-wide significant loci that displayed directionally discordant effects in the two degree of freedom DINGO test exhibited p-values that were far less extreme using the simple meta-analytic strategy, including some loci that may have been missed entirely (e.g. the *GLIS3* locus discussed below). The implication is that a two degree of freedom test may be the preferable strategy in the case of phenotypes that are known to exhibit substantial numbers of loci with directionally discordant effects (e.g. perinatal growth-related traits like birth weight), whereas a simple meta-analytic strategy may be the superior strategy in the case of other traits (bearing in mind that such knowledge about likely underlying genetic architecture may not always be available from previous GWAS a *priori*). We have implemented both the meta-analytic tests and the two degree of freedom test as part of the DINGO package in the publicly available web-based software CTG-VL^17^.

We would like to highlight two findings from our empirical GWAS of birth weight that we think deserve further attention. The first is the *GLIS3* locus. As mentioned above, it is unlikely that this locus would have been flagged by researchers adopting a simple meta-analytic strategy, although interestingly the locus was identified previously in the deCODE paper using separate maternal and fetal GWAS (although not discussed explicitly by the authors). *GLIS3* encodes a member of the GLI-similar zinc finger protein family. The GLIS3 protein functions as both a repressor and activator of transcription that is involved in the development of pancreatic beta cells. Homozygous mutant *Glis3* mice develop neonatal diabetes due to insufficient pancreatic beta cells ^22^, and deletions of the gene in humans are associated with neonatal diabetes ^23-25^. The “A” allele of the rs7034200 variant within the *GLIS3* gene (i.e. which in our study is associated with increased offspring birth weight when present in the maternal genome, but decreased birth weight when present in the fetal genome), has also been associated with increased risk of type 2 diabetes ^26-31^, higher fasting glucose levels ^28,32,33^ and impaired beta cell function ^28,33-35^. The glucose raising allele at this SNP is also strongly associated with impaired beta-cell function in non-diabetic adults ^33,35^ and in healthy children and adolescents ^32^. We therefore hypothesise that diabetes predisposing variants at this locus may increase levels of circulating glucose in pregnant mothers (i.e. which would tend to increase intrauterine growth and offspring birth weight), but simultaneously act to decrease offspring birth weight when the same alleles are transmitted to the fetus (e.g. by impairing beta cell function and therefore fetal insulin secretion). Similar mechanisms are thought to underlie other diabetes predisposing variants that show directionally inconsistent maternal and fetal genetic associations with birth weight^6,7,36^.

The second result we would like to highlight is the genome-wide significant SNP rs11085720 in an intron in the *DNMT1* gene. Although this SNP was not significant in the deCODE meta-analysis, it was strongly significant in both the two degree of freedom (p-value = 8.6 × 10^−13^) and one degree of freedom meta-analyses (p_min_ = 2.3 × 10^−12^). The gene *DNMT1* codes for the enzyme DNA methyltransferase 1 which is responsible for the addition of methyl groups to specific CpG structures in DNA. Methylation of CpG islands is associated with transcriptional silencing and knockout experiments suggest that this enzyme is responsible for the bulk of methylation in mouse cells and is essential for embryonic development^37^. In humans, rare variants in the *DNMT1* gene are associated with forms of cerebellar ataxia^38^ and neuropathy^39,40^. Although we cannot be sure that *DNMT1* is indeed the causative gene at this locus, we highlight its potential involvement because of the considerable interest in the relationship between adverse environmental exposures during pregnancy, possible mediation via DNA methylation and long-term effects on disease risk due to putative intrauterine programming. In this respect, it is interesting to note that conditional analyses suggest that this locus involves both maternal and fetal effects. How methylation levels in mothers could in turn affect the birth weight of their offspring is unclear, however it is interesting that studies of knock-out mice have shown that a lack of both maternal and zygotic *Dnmt1* results in complete demethylation of imprinted genes in (mouse) blastocysts^41^.

Several simple extensions to/applications of our methods present themselves. First, although we have applied our methods to birth weight, there exists many other traits where maternal and offspring GWAS have been conducted including gestational duration^9,10,42^ and placental weight^43^ which involve only partially overlapping samples and could benefit from reanalysis using the framework that we have outlined in this manuscript. Second, there is increasing interest in and growing realization in the scientific community that indirect genetic effects contribute not only to perinatal traits like birth weight^1,6,7,18,44^, but also to many later life socially patterned traits like educational attainment^14-16^. The implication is that substantial gains in power to detect loci underlying many of these traits might be achieved by combining parental and offspring GWAS either through a simple meta-analytic strategy, or by performing multi degree of freedom tests across the genome to GWAS summary results data derived from parents and children. Third, although the development of DINGO was motivated by the analysis of traits that are affected by indirect maternal/paternal effects, there is no reason why analogous methods cannot also be applied to traits where e.g. the offspring’s genome affects e.g. the mother’s phenotype. For example, offspring genotype is thought to affect risk of maternal preeclampsia and so it would be possible in theory to combine a fetal GWAS of preeclampsia with a GWAS of maternal preeclampsia using a similar procedure for binary traits^45,46^ (see below). Other phenotypes such as twinning and fertility/fecundity might also be interesting candidates for such analyses. Fourth, we note that our framework can be easily extended to a three degree of freedom test that incorporates summary results statistics from maternal, paternal and “fetal” GWAS (see **Supplementary Note**). Whilst a three degree of freedom test would most likely be inefficient for the analysis of perinatal traits (i.e. where indirect paternal genetic effects are likely to be negligible and so the estimation of a paternal genetic effect parameter would be of little benefit whilst requiring an extra degree of freedom), such a test could potentially be useful for the analysis of traits like educational attainment which are known to involve all three sorts of effect ^16^. Alternatively, employing a simple meta-analytic approach, but combining across maternal, paternal and offspring GWAS could also be advantageous (also see **Supplementary Note**).

There are several limitations to the methods proposed in this manuscript. First, the two degree of freedom test and the meta-analytic strategy utilize LD score regression to provide an accurate estimate of effective sample overlap. Therefore, all factors discussed in the literature that may affect the accuracy/precision of the LD score intercept apply *mutatis mutandis* to the present methods. We recommend that the sample sizes of both maternal and fetal GWAS be large and as ancestrally homogenous as possible to accurately estimate sample overlap. Second, the formulae for the two degree of freedom and meta-analytic tests that we have derived requires that the coefficients from the maternal and fetal GWAS are from a linear regression analysis of quantitative traits. It is critically important that these different GWAS are analysed on (or can be transformed to) the same scale. In the case of binary traits, a number of approaches exist for transforming logistic regression coefficients to a (normal) liability scale^47-50^. In theory it should be possible to transform these logistic coefficients (and their standard errors) to the liability scale and then apply the methods outlined in this manuscript to permit a similar analysis of binary traits. We expect that analogous to quantitative traits, such a method would improve the power to detect loci that simultaneously exert direct and indirect genetic effects and would accurately model their contribution to trait variation through maternal and fetal pathways. In contrast, existing methods for analysing binary data are geared towards increasing the power to detect direct genetic effects only through the utilization of self-report data from ungenotyped related individuals, or are appropriate for the analysis of individual level genotypes (i.e. rather than summary results data)^51-53^. Finally, our power simulations and empirical analyses used a Bonferroni correction to determine the genome-wide significance of the one degree of freedom tests. Simulations we have conducted suggest that this correction is appropriate although very slightly conservative in the case of substantial sample overlap and phenotypic correlation (see **Supplementary Note**). Thus, our analyses may have slightly underestimated the true power of the one degree of freedom tests under these conditions.

In conclusion, in this manuscript we have introduced two new methods designed to increase the power of locus discovery in meta-analyses of summary results statistics from maternal and fetal GWAS. We have shown through a combination of analytical results, simulation and empirical analysis that a simple one degree of freedom meta-analytic strategy where signals from maternal and fetal GWAS are meta-analysed together conditional on the estimated sample overlap is likely to be powerful strategy in the analysis of perinatal traits. Indeed, adopting this strategy increased the number of known birth weight loci by 62 and implicated a number of known and novel pathways in the aetiology of the trait. We have also shown that a two degree of freedom test may be particularly powerful strategy for analysing traits where a substantial proportion of loci involve discordant maternal and fetal genetic effects of similar magnitude. The two degree of freedom test and the one degree of freedom meta-analytic procedure described in this manuscript are implemented in the “<u>D</u>irect and <u>IN</u>direct effects analysis of <u>G</u>enetic l<u>O</u>ci” (DINGO) package, available as part of the CTG-VL software.^17^

## Data Availability

All data produced in the present study are available upon reasonable request to the authors.

https://www.decode.com/summarydata/

https://www.ukbiobank.ac.uk/

## Acknowledgements

This work was supported by Australian National Health and Medical Research Council (NHMRC) grants (GNT1183074, GNT1157714). D.M.E. is supported by an NHMRC Leadership Fellowship (2017942). N.M.W is supported by an NHMRC Emerging Leadership Fellowship (2008723). G.H.M. is the recipient of an Australian Research Council Discovery Early Career Award (Project number: DE220101226) funded by the Australian Government and supported by the Research Council of Norway (Project grant: 325640). L.Y. is supported by a Australian Research Council Future Fellowship (FT220100069). R.M.F. and R.N.B. are supported by a Wellcome Senior Research Fellowship (WT220390). R.M.F. is also supported by a grant from the Eunice Kennedy Shriver National Institute Of Child Health & Human Development of the National Institutes of Health under Award Number R01HD101669. This research was funded in part by the Wellcome Trust (WT220390). Human genotype and phenotype data from the UKB on which the results of simulations were based were accessed with accession ID 53641.

