## Supplementary Figure for "Direct and INdirect effects analysis of Genetic lOci (DINGO): A software package to increase the power of locus discovery in GWAS meta-analyses of perinatal phenotypes and traits influenced by indirect genetic effects"

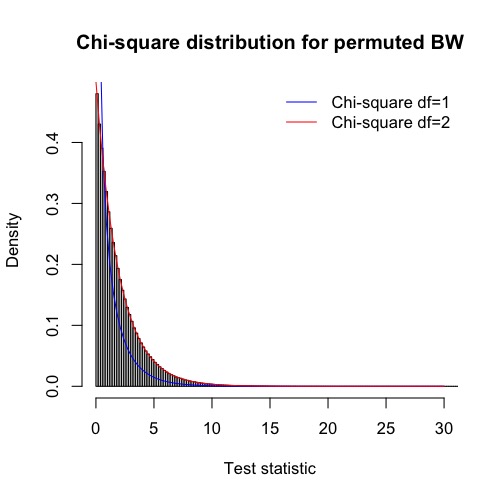

**Supplementary Figure 1. Empirical distribution of the two degree of freedom test statistic *T_2df_* across the genome under a single permuted dataset in the UK Biobank.** Summary results statistics were thinned by extracting one in every thousand SNPs (i.e. to correct for dependencies that might arise through linkage disequilibrium at close by markers) and checked against known distributions using the one-sample Kolmogorov-Smirnov test. Blue and red lines indicate the expected probability density for a chi-square 1 and chi-square 2 distribution respectively. The empirical distribution closely matched the two degree of freedom chi-square distribution (Kolmogorov-Smirnov p-value = 0.86).

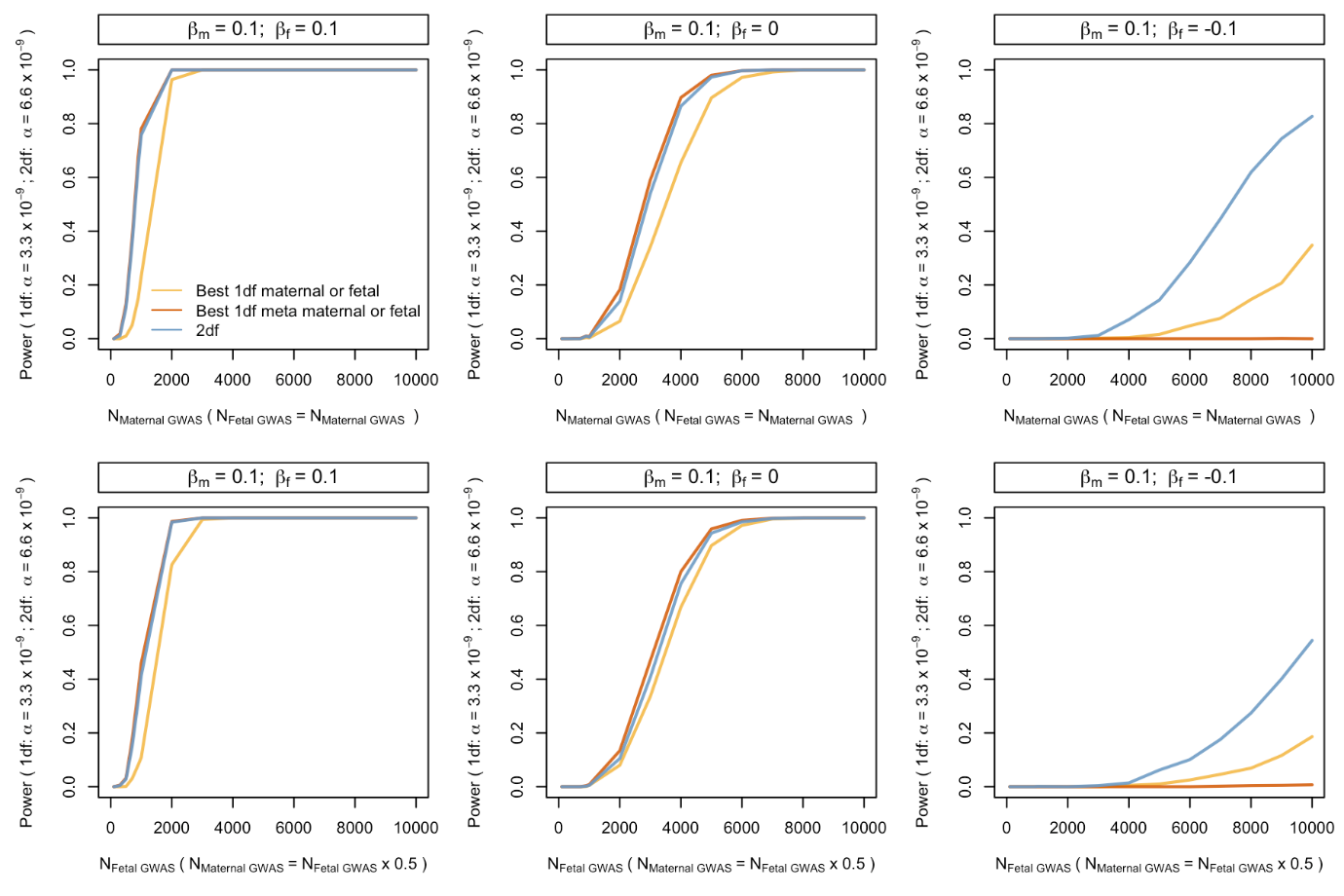

**Supplementary Figure 2.** **Power to detect association (as evaluated using simulation) using a traditional strategy of analysing separate maternal and fetal GWAS, a strategy involving one degree of freedom meta-analyses, and a strategy of performing two degree of freedom *T_2df_* tests across the genome. The effect of varying the sample size of the maternal GWAS is shown.** β_m_ and β_f_ refer to maternal and fetal genetic effects on a standardized trait. Data were simulated assuming no sample overlap. In the top row, the sample sizes for maternal GWAS and fetal GWAS are the same. In the bottom row, the sample size for the maternal GWAS is half the fetal GWAS. For the traditional strategy of running separate maternal and fetal GWAS and the one degree of freedom meta-analysis strategy, we set the alpha value to α = 3.3 x 10^-9^ i.e. half the α = 6.6 x 10^-9^ type I error rate of the two degree of freedom *T_2df_* test in order to take into account that we are performing twice the number of statistical tests in the former situations. In the case of the traditional strategy of running separate maternal and fetal GWAS and the one degree of freedom meta-analysis strategy, we evaluated power with respect to whether *either* test met the criterion for genome-wide significance for each replicate.

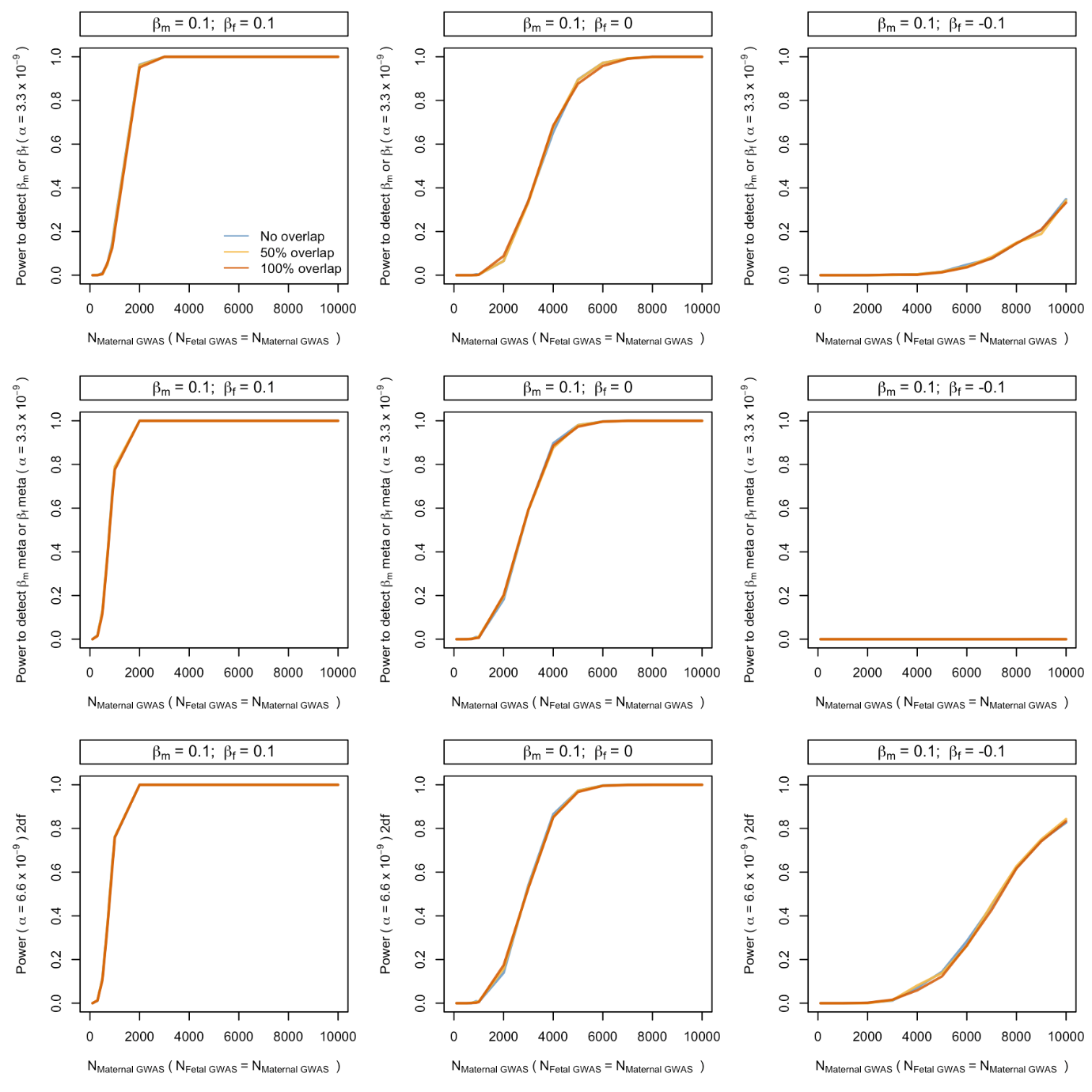

**Supplementary Figure 3.** **Power to detect association (as evaluated using simulation) using a traditional strategy of analysing separate maternal and fetal GWAS (top panel), a strategy involving one degree of freedom meta-analyses (middle panel), and a strategy of performing two degree of freedom *T_2df_* tests across the genome (bottom panel). The effect of varying the degree of sample overlap is shown.** β_m_ and β_f_ refer to maternal and fetal genetic effects on a standardized trait. Data were simulated assuming no residual correlation between maternal and offspring phenotypes (ρ = 0). For the traditional strategy of running separate maternal and fetal GWAS and the one degree of freedom meta-analysis strategy, we set the alpha value to α = 3.3 x 10^-9^ i.e. half the α = 6.6 x 10^-9^ type I error rate of the two degree of freedom *T_2df_* test in order to take into account that we are performing twice the number of statistical tests in the former situations. In the case of the traditional strategy of running separate maternal and fetal GWAS and the one degree of freedom meta-analysis strategy, we evaluated power with respect to whether *either* test met the criterion for genome-wide significance for each replicate.

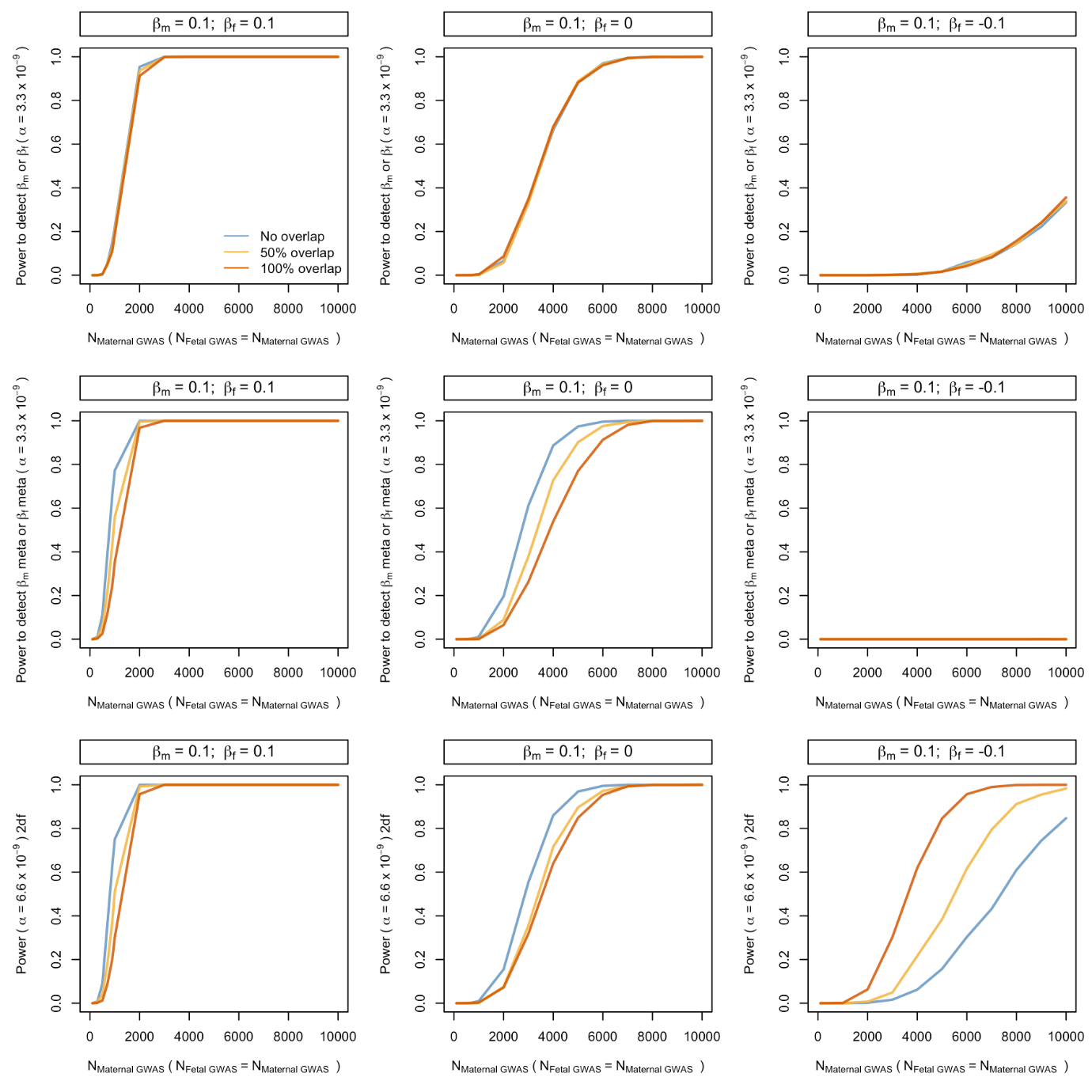

**Supplementary Figure 4.** **Power to detect association (as evaluated using simulation) using a traditional strategy of analysing separate maternal and fetal GWAS (top panel), a strategy involving one degree of freedom meta-analyses (middle panel), and a strategy of performing two degree of freedom *T_2df_* tests across the genome (bottom panel). The effect of varying the degree of sample overlap is shown.** β_m_ and β_f_ refer to maternal and fetal genetic effects on a standardized trait. Data were simulated assuming high residual correlation between maternal and offspring phenotypes (ρ = 0.5). For the traditional strategy of running separate maternal and fetal GWAS and the one degree of freedom meta-analysis strategy, we set the alpha value to α = 3.3.x10^-9^ i.e. half the α = 6.6.x10^-9^ type I error rate of the two degree of freedom *T_2df_* test in order to take into account that we are performing twice the number of statistical tests in the former situations. In the case of the traditional strategy of running separate maternal and fetal GWAS and the one degree of freedom meta-analysis strategy, we evaluated power with respect to whether *either* test met the criterion for genome-wide significance for each replicate.

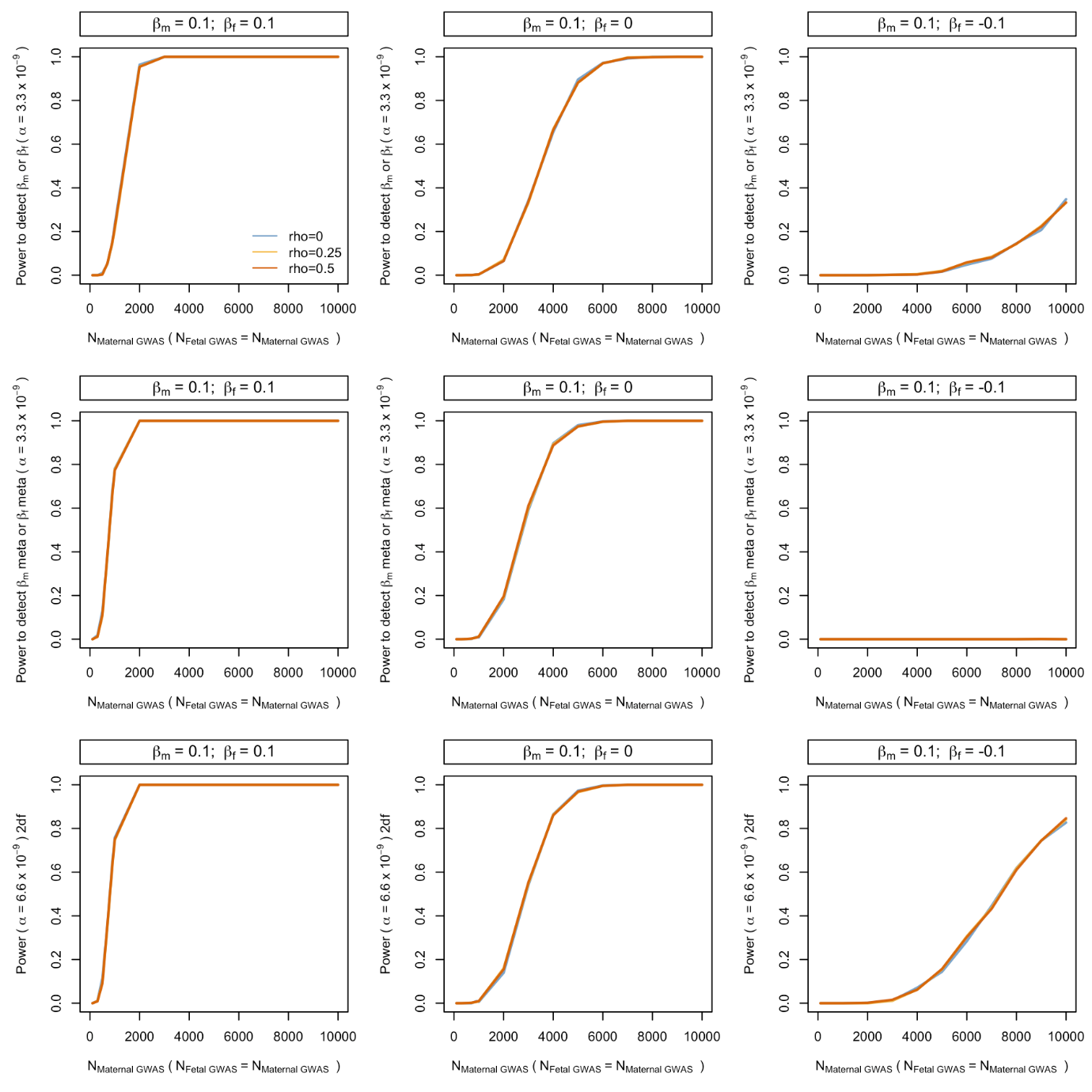

**Supplementary Figure 5.** **Power to detect association (as evaluated using simulation) using a traditional strategy of analysing separate maternal and fetal GWAS (top panel), a strategy involving one degree of freedom meta-analyses (middle panel), and a strategy of performing two degree of freedom *T_2df_* tests across the genome (bottom panel). The effect of varying the residual phenotypic correlation (****ρ) is shown. Data were simulated assuming no sample overlap between maternal and fetal GWAS.** β_m_ and β_f_ refer to maternal and fetal genetic effects on a standardized trait. For the traditional strategy of running separate maternal and fetal GWAS and the one degree of freedom meta-analysis strategy, we set the alpha value to α = 3.3 x 10^-9^ i.e. half the α = 6.6 x 10^-9^ type I error rate of the two degree of freedom *T_2df_* test in order to take into account that we are performing twice the number of statistical tests in the former situations. In the case of the traditional strategy of running separate maternal and fetal GWAS and the one degree of freedom meta-analysis strategy, we evaluated power with respect to whether *either* test met the criterion for genome-wide significance for each replicate.

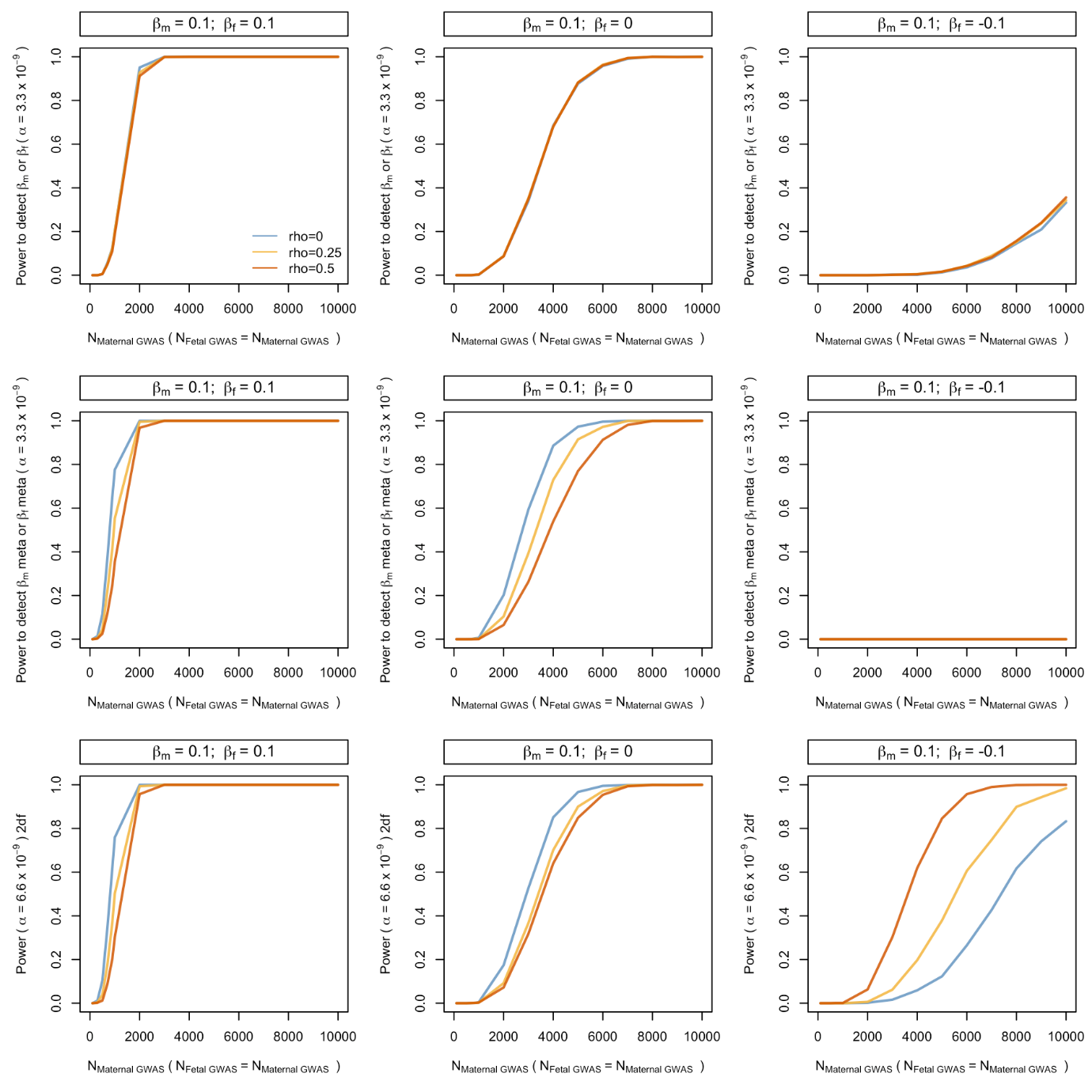

**Supplementary Figure 6.** **Power to detect association (as evaluated using simulation) using a traditional strategy of analysing separate maternal and fetal GWAS (top panel), a strategy involving one degree of freedom meta-analyses (middle panel), and a strategy of performing two degree of freedom *T_2df_* tests across the genome (bottom panel). The effect of varying the residual phenotypic correlation (ρ) is shown. Data were simulated assuming no sample overlap between maternal and fetal GWAS.** β_m_ and β_f_ refer to maternal and fetal genetic effects on a standardized trait. For the traditional strategy of running separate maternal and fetal GWAS and the one degree of freedom meta-analysis strategy, we set the alpha value to α = 3.3 x 10^-9^ i.e. half the α = 6.6 x 10^-9^ type I error rate of the two degree of freedom *T_2df_* test in order to take into account that we are performing twice the number of statistical tests in the former situations. In the case of the traditional strategy of running separate maternal and fetal GWAS and the one degree of freedom meta-analysis strategy, we evaluated power with respect to whether *either* test met the criterion for genome-wide significance for each replicate.

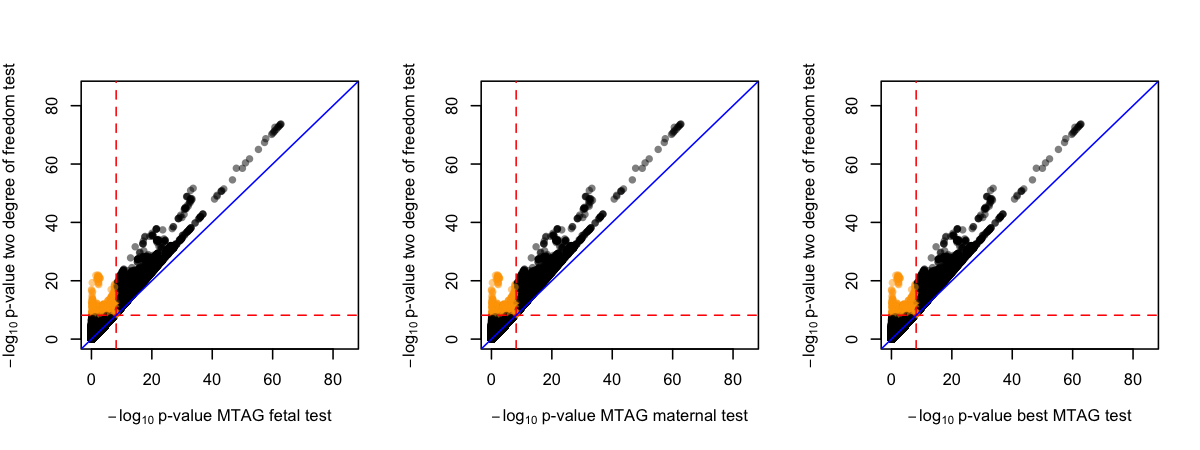

**Supplementary Figure 7. P-value scatter plots comparing MTAG results versus the two degree of freedom test in the GWAS analysis of birth weight.** The -log_10_p-value from the two degree of freedom test was compared against the -log_10_p-value of SNPs from (left) the MTAG results for own birth weight, (middle) the MTAG results for offspring birth weight, and (right) the strongest MTAG p-value at the locus. Red dashed lines denote the genome-wide significant thresholds of α = 3.3 x 10^-9^ for the one degree of freedom MTAG analyses and α = 6.6 x 10^-9^ for the two degree of freedom test. Blue diagonal lines indicate x=y. Orange circles are SNPs that are only significant using the two degree of freedom test.
