## Supplementary Note for "Direct and INdirect effects analysis of Genetic lOci (DINGO): A software package to increase the power of locus discovery in GWAS meta-analyses of perinatal phenotypes and traits influenced by indirect genetic effects"

For the derivations below, we assume that the population model for the phenotype is given by the linear function:

$$y_{i}=\alpha+\beta_{f}x_{f_{i}}+\beta_{m}x_{m_{i}}+u_{i}$$

where $y$ is the phenotype to be modelled, $x_{f}$ and $x_{m}$ are the dosage of an individual’s own (i.e. fetal) and their mother’s SNP (i.e. maternal) respectively, $\beta_{f}$ and $\beta_{m}$ are the true population fetal and maternal genetic effects on the phenotype at these SNPs, $\alpha$ is an intercept, $u$ is an error term that is mean independent of the SNP dosages i.e. $E\left( {u|x}_{f},x_{m} \right)=0$ and represents the sum total of all unmodelled influences on $y$, and the *i* subscript indexes individual *i*.

***Regression coefficients from maternal and fetal GWAS are biased and inconsistent in the presence of both maternal and fetal genetic effects at the same locus***

The simple regression of an individual’s own phenotype ($y$) on their own SNP dosage (*x_f_*) is an inconsistent estimator of the true direct fetal genetic effect ($\beta_{f}$) in the presence of maternal genetic effects ($\beta_{m}$) at the same locus i.e.:

$\mathrm{plim}(\hat{b}_{f})=plim\left( \frac{\mathrm{cov}\left( y,x_{f} \right)}{\mathrm{var}\left( x_{f} \right)} \right)=\frac{\mathrm{COV}\left( y, x_{f} \right)}{\mathrm{VAR}\left( x_{f} \right)}=\frac{\mathrm{VAR}\left( x_{f} \right)\left( \beta_{f}+\frac{1}{2}\beta_{m} \right)}{\mathrm{VAR}\left( x_{f} \right)}=$ $\beta_{f}+\frac{1}{2}\beta_{m}$

where cov and var refers to the sample covariance and variance, COV and VAR the population variance and covariance, and plim is the probability limit.

Likewise, the simple regression of an individual’s own phenotype ($y_{i}$) on their own SNP dosage ($x_{f_{i}}$) is a biased estimator of the true direct fetal genetic effect ($\beta_{f}$) in the presence of indirect maternal genetic effects ($\beta_{m}$) at the same locus i.e.:

$$E(\hat{b}_{f})= {E(E(\hat{\beta}}_{f_{OLS}}|\boldsymbol{x}_{\boldsymbol{f}}))$$

$$=E(E( \frac{\sum_{i=1}^{N} \left( x_{f_{i}}-\bar{x}_{f} \right)y_{i}}{\sum_{i=1}^{N} \left( x_{f_{i}}-\bar{x}_{f} \right)x_{f_{i}}}|\boldsymbol{x}_{\boldsymbol{f}}))$$

$$=E(E( \frac{\sum_{i=1}^{N} \left( x_{f_{i}}-\bar{x}_{f} \right)\left( \alpha+{\beta_{f}x}_{f_{i}}+{\beta_{m}x}_{m_{i}}+u_{i} \right)}{\sum_{i=1}^{N} \left( x_{f_{i}}-\bar{x}_{f} \right)x_{f_{i}}}|\boldsymbol{x}_{\boldsymbol{f}}))$$

$$=E(E( \frac{\alpha\sum_{i=1}^{N} \left( x_{f_{i}}-\bar{x}_{f} \right)}{\sum_{i=1}^{N} \left( x_{f_{i}}-\bar{x}_{f} \right)x_{f_{i}}}+\frac{\beta_{f}\sum_{i=1}^{N} \left( x_{f_{i}}-\bar{x}_{f} \right)x_{f_{i}}}{\sum_{i=1}^{N} \left( x_{f_{i}}-\bar{x}_{f} \right)x_{f_{i}}}+\frac{\beta_{m}\sum_{i=1}^{N} \left( x_{f_{i}}-\bar{x}_{f} \right)x_{m_{i}}}{\sum_{i=1}^{N} \left( x_{f_{i}}-\bar{x}_{f} \right)x_{f_{i}}}+\frac{\sum_{i=1}^{N} \left( x_{f_{i}}-\bar{x}_{f} \right)u_{i}}{\sum_{i=1}^{N} \left( x_{f_{i}}-\bar{x}_{f} \right)x_{f_{i}}}|\boldsymbol{x}_{\boldsymbol{f}}))$$

$$=0+\beta_{f}+\beta_{m}\frac{\mathrm{cov}(x_{f},x_{m})}{\mathrm{var}(x_{f})}+\frac{\sum_{i=1}^{N} \left( x_{f_{i}}-\bar{x}_{f} \right)}{\sum_{i=1}^{N} \left( x_{f_{i}}-\bar{x}_{f} \right)x_{f_{i}}}E(E( u_{i}|\boldsymbol{x}_{\boldsymbol{f}}))$$

$$=\beta_{f}+\beta_{m}\frac{\mathrm{cov}(x_{f},x_{m})}{\mathrm{var}(x_{f})}$$

$=\beta_{f}+{\frac{1}{2}\beta}_{m}$ in large samples.

By a similar process it can be shown that the simple regression of offspring phenotype on maternal SNP is a biased and inconsistent estimate of the true indirect maternal genetic effect ($\beta_{m}$) in the presence of direct fetal effects ($\beta_{f}$) at the same locus i.e.:

$\mathrm{plim}(\hat{b}_{m})=$ $\beta_{m}+\frac{1}{2}\beta_{f}$

$E(\hat{b}_{m})=$ $\beta_{m}+\beta_{f}\frac{\mathrm{cov}(x_{f},x_{m})}{\mathrm{var}(x_{m})}= \beta_{m}+{\frac{1}{2}\beta}_{f}$ in large samples.

***Asymptotic distribution of the two degree of freedom test T_2df_ under the null and alternative hypotheses***

The joint sampling distribution of maternal and fetal effect estimates is bivariate normal in large samples:

$$\binom{\hat{\beta}_{f}}{\hat{\beta}_{m}}\sim N\left( \binom{\beta_{f}}{\beta_{m}} , \boldsymbol{\Sigma} \right)$$

with $\boldsymbol{\Sigma}=\left( \begin{matrix} \mathrm{var}(\hat{\beta}_{f}) & \mathrm{cov}(\hat{\beta}_{f},\hat{\beta}_{m}) \\ \mathrm{cov}(\hat{\beta}_{f},\hat{\beta}_{m}) & \mathrm{var}(\hat{\beta}_{m}) \end{matrix} \right)$

The T*_2df_* test statistic is given by:

$$T_{2df}=\left( \hat{\beta}_{f} \right.\left. \hat{\beta}_{m} \right)\left( \begin{matrix} \mathrm{var}(\hat{\beta}_{f}) & \mathrm{cov}(\hat{\beta}_{f},\hat{\beta}_{m}) \\ \mathrm{cov}(\hat{\beta}_{f},\hat{\beta}_{m}) & \mathrm{var}(\hat{\beta}_{m}) \end{matrix} \right)^{-1}\binom{\hat{\beta}_{f}}{\hat{\beta}_{m}}=\mathbf{b}\mathbf{'}\boldsymbol{\Sigma}^{\boldsymbol{-}\boldsymbol{1}}\mathbf{b}$$

Let

$$\mathbf{S=}\begin{matrix} \mathrm{SE}(\hat{\beta}_{f}) & 0 \\ 0 & \mathrm{SE}(\hat{\beta}_{m}) \end{matrix}$$

We can therefore rewrite T*_2df_* as:

$$T_{2df}=\mathbf{b}\mathbf{'}{\mathbf{S}^{\boldsymbol{-}\boldsymbol{1}\mathbf{'}}\mathbf{S}\mathbf{'}\boldsymbol{\Sigma}}^{\boldsymbol{-}\boldsymbol{1}}\mathbf{S}\mathbf{S}^{\boldsymbol{-}\boldsymbol{1}}\mathbf{b}\boldsymbol{=}\mathbf{z}\mathbf{'}\mathbf{R}^{\boldsymbol{-}\boldsymbol{1}}\mathbf{z}$$

where $\boldsymbol{z\sim}N\boldsymbol{(\mu,R)}$ and $\mathbf{R}$ is the correlation matrix of fetal and maternal effect estimates.

Let $\boldsymbol{\Lambda}$ and **U** be the diagonal matrix of eigenvalues and the orthogonal matrix of eigenvectors of $\mathbf{R}$ respectively. Performing a eigen decomposition on $\mathbf{R}$ and inverting we have:

$${\mathbf{R}^{\boldsymbol{-}\boldsymbol{1}}\mathbf{=}\boldsymbol{U\Lambda}}^{\boldsymbol{-}\boldsymbol{1}}\mathbf{U}\mathbf{'}$$

If we let $\boldsymbol{w=\Lambda}^{\boldsymbol{-}\frac{\boldsymbol{1}}{\boldsymbol{2}}}\mathbf{U'z}$, then

$${T_{2df}=\mathbf{z}}^{\mathbf{'}}\mathbf{U}\boldsymbol{\Lambda}^{\boldsymbol{-}\boldsymbol{1}}\mathbf{U}^{\boldsymbol{'}}\mathbf{z}\mathbf{=(}\mathbf{z}\mathbf{'}\mathbf{U}\boldsymbol{\Lambda}^{\boldsymbol{-}\frac{\boldsymbol{1}}{\boldsymbol{2}}}\boldsymbol{)}{\mathbf{(}\boldsymbol{\Lambda}}^{\boldsymbol{-}\frac{\boldsymbol{1}}{\boldsymbol{2}}}\mathbf{U}^{\boldsymbol{'}}\mathbf{z}\mathbf{)=}\mathbf{w}\mathbf{'}\mathbf{w}$$

with E(**w**) = $\boldsymbol{\Lambda}^{\boldsymbol{-}\frac{\boldsymbol{1}}{\boldsymbol{2}}}\mathbf{U'}\boldsymbol{\mu}$ and Var(**w**) = $\boldsymbol{\Lambda}^{\boldsymbol{-}\frac{\boldsymbol{1}}{\boldsymbol{2}}}\mathbf{U}^{\mathbf{'}}\mathbf{RU}\boldsymbol{\Lambda}^{\boldsymbol{-}\frac{\boldsymbol{1}}{\boldsymbol{2}}}\boldsymbol{=}\boldsymbol{\Lambda}^{\boldsymbol{-}\frac{\boldsymbol{1}}{\boldsymbol{2}}}\mathbf{U}^{\mathbf{'}}\boldsymbol{U\Lambda}\mathbf{U}^{\mathbf{'}}\mathbf{U}\boldsymbol{\Lambda}^{\boldsymbol{-}\frac{\boldsymbol{1}}{\boldsymbol{2}}}\boldsymbol{=}\mathbf{I}$, where

$$\boldsymbol{w\sim}N(\boldsymbol{\Lambda}^{\boldsymbol{-}\frac{\boldsymbol{1}}{\boldsymbol{2}}}\mathbf{U'}\boldsymbol{\mu},\mathbf{I})$$

Under the null hypothesis of no maternal and fetal effect at the variant being tested, $\boldsymbol{\mu=0}$. In this case, **w** is a 2 × 1 vector of standard normal variables. Thus, under H_0_ the T*_2df_* test statistic follows a central chi-squared distribution with two degrees of freedom:

$$T_{2df}\sim\chi_{2}^{2}$$

Under the alternative hypothesis that there exists a maternal and fetal effect of the SNP, $\boldsymbol{\mu\neq0}$. Let $\boldsymbol{\eta=}\boldsymbol{\Lambda}^{\boldsymbol{-}\frac{\boldsymbol{1}}{\boldsymbol{2}}}\mathbf{U'}\boldsymbol{\mu}$, then $\boldsymbol{w\sim}N(\boldsymbol{\eta},\mathbf{I})$. The sum of squares of **w** follows a non-central chi-square distribution with two degrees of freedom and non-centrality parameter NCP = $\boldsymbol{\eta}^{\boldsymbol{'}}\boldsymbol{\eta=}{\boldsymbol{\mu}^{\boldsymbol{'}}\mathbf{U}\boldsymbol{\Lambda}^{\boldsymbol{-}\frac{\boldsymbol{1}}{\boldsymbol{2}}}\boldsymbol{\Lambda}}^{\boldsymbol{-}\frac{\boldsymbol{1}}{\boldsymbol{2}}}\mathbf{U}^{\mathbf{'}}\boldsymbol{\mu=}\boldsymbol{\mu}^{\boldsymbol{'}}\boldsymbol{\Sigma}^{\boldsymbol{-1}}\boldsymbol{\mu}$. It follows that under H_1_:

$$T_{2df}\sim\chi_{2}^{2}(\boldsymbol{\mu}^{\boldsymbol{'}}\boldsymbol{\Sigma}^{\boldsymbol{-}\boldsymbol{1}}\boldsymbol{\mu})$$

***Distribution of the two degree of freedom test statistic T_2df_ under the Null Hypothesis of no association between SNPs and trait***

To validate whether our test statistic was distributed as a two degree of freedom chi-square test under the Null Hypothesis of no association, we applied our model to a single permuted dataset in the UK Biobank. We extracted phenotypes of birth weight and offspring birth weight from the UK Biobank. Exclusion criteria included multiple births, inconsistent reports of birth weight between time points, birth weight >4.5 kilograms or <2.5 kilograms, withdrawal from the UK Biobank, and non-European ancestry. Only unrelated individuals, defined as having genomic similarity less than 3^rd^ degree relatives^1^, were included in the final sample. This resulted in 118,016 individuals with own birth weight only, 91,164 mothers with offspring birth weight only, and 103,701 mothers with both own and offspring birth weight. Own birth weight was regressed on sex and assessment centre (offspring birth weight was regressed on assessment centre only since sex was not available), and the residuals were then transformed into z-scores. See the Warrington et al. (2019)^2^ paper for details of phenotype preparation. We then permuted individuals’ phenotypes with respect to their genotypes (i.e. individual’s own birth weight and offspring birth weight were permuted together when both measures were present). We performed fetal GWAS on own birth weight and maternal GWAS on offspring birth weight using fastGWA^3^. We then applied the two degree of freedom test to the regression coefficients and their standard errors from the two GWAS for each SNP across the genome. Given that mothers with own and offspring birth weight were included in both GWAS, we used LD score regression to estimate the degree of sample overlap (LD score intercept = 0.1192). Lastly, we thinned the summary results statistics by extracting one in every thousand results (i.e. to correct for dependencies that might arise through linkage disequilibrium at close by markers) checked the distribution of test statistics using the one-sample Kolmogorov-Smirnov test.

The empirical distribution of the test statistics under the Null Hypothesis of no association are shown in Supplementary Figure 1 and closely match a two degree of freedom chi-square distribution (Kolmogorov-Smirnov p-value = 0.86).

The UK Biobank study was approved by the UK National Health Service National Research Ethics Service. Written consent was obtained from both the participants and their parents (for subjects younger than 18 years old). This study was approved by the Human Research Ethics Committee at the University of Queensland (approval number: 2019002705).

***Derivation of the three degree of freedom test***

We assume that the population model for the phenotype is given by the linear function:

$$y=\alpha+\beta_{f}x_{f}+\beta_{m}x_{m}+\beta_{p}x_{p}+u$$

where $y$ is the phenotype to be modelled, $x_{f}$ , $x_{m}$ and $x_{p}$ are the dosage of fetal and maternal SNPs respectively, $\beta_{f}$ $\beta_{m}$ and $\beta_{p}$ and are the true population fetal, maternal and paternal genetic effects on the phenotype at these SNPs, $\alpha$ is an intercept, and $u$ is an error term that is mean independent of the SNP dosages i.e. $E\left( {u|x}_{f},x_{m},x_{p} \right)=0$ and represents the sum total of all unmodelled influences on y.

We can estimate the true direct fetal ($\beta_{f}$) and indirect maternal ($\beta_{m}$) and paternal ($\beta_{p}$) genetic effects using the following weighted linear combination of regression coefficients derived from empirical linear regressions of phenotype on fetal SNP ($\hat{b}_{f}$), maternal SNP ($\hat{b}_{m}$) and paternal SNP ($\hat{b}_{p}$):

$$\hat{\beta}_{f}=2\hat{b}_{f}-\hat{b}_{m}-\hat{b}_{p}$$

$$\hat{\beta}_{m}=\frac{3}{2}\hat{b}_{m}-\hat{b}_{f}+\frac{1}{2}\hat{b}_{p}$$

$$\hat{\beta}_{p}=\frac{3}{2}\hat{b}_{p}-\hat{b}_{f}+\frac{1}{2}\hat{b}_{m}$$

and their standard errors:

$${\mathrm{SE}(\hat{\beta}}_{f})=\sqrt{4\mathrm{var}\left( \hat{b}_{f} \right)+{\mathrm{var}(\hat{b}}_{m})+\mathrm{var}\left( \hat{b}_{p} \right)+2\mathrm{cov}\left( \hat{b}_{m},\hat{b}_{p} \right)-4\mathrm{cov}\left( \hat{b}_{f},\hat{b}_{m} \right)-4\mathrm{cov}\left( \hat{b}_{f},\hat{b}_{p} \right)}$$

$$=\sqrt{4\mathrm{var}\left( \hat{b}_{f} \right)+{\mathrm{var}(\hat{b}}_{m})+\mathrm{var}\left( \hat{b}_{p} \right)+2\times\mathrm{int}_{m.p}\times SE\left( \hat{b}_{m} \right)\mathrm{SE}\left( \hat{b}_{p} \right)-4\times\mathrm{int}_{f,m}\times SE\left( \hat{b}_{f} \right)\mathrm{SE}(\hat{b}_{m})-4\times\mathrm{int}_{f,p}\times SE\left( \hat{b}_{f} \right)\mathrm{SE}(\hat{b}_{p})}$$

$${\mathrm{SE}(\hat{\beta}}_{m})=\sqrt{\frac{9}{4}\mathrm{var}\left( \hat{b}_{m} \right)+{\mathrm{var}(\hat{b}}_{f})+\frac{1}{4}\mathrm{var}\left( \hat{b}_{p} \right)-\mathrm{cov}\left( \hat{b}_{f},\hat{b}_{p} \right)-3\mathrm{cov}\left( \hat{b}_{f},\hat{b}_{m} \right)+\frac{3}{2}\mathrm{cov}\left( \hat{b}_{m},\hat{b}_{p} \right)}$$

$$=\sqrt{\frac{9}{4}\mathrm{var}\left( \hat{b}_{m} \right)+{\mathrm{var}(\hat{b}}_{f})+\frac{1}{4}\mathrm{var}\left( \hat{b}_{p} \right)-\mathrm{int}_{f,p}\times SE\left( \hat{b}_{f} \right)\mathrm{SE}\left( \hat{b}_{p} \right)-3\times\mathrm{int}_{f,m}\times SE\left( \hat{b}_{f} \right)\mathrm{SE}\left( \hat{b}_{m} \right)+\frac{3}{2}\times\mathrm{int}_{m,p}\times SE\left( \hat{b}_{m} \right)\mathrm{SE}(\hat{b}_{p})}$$

$${\mathrm{SE}(\hat{\beta}}_{p})=\sqrt{\frac{9}{4}\mathrm{var}\left( \hat{b}_{p} \right)+{\mathrm{var}(\hat{b}}_{f})+\frac{1}{4}\mathrm{var}\left( \hat{b}_{m} \right)-\mathrm{cov}\left( \hat{b}_{f},\hat{b}_{m} \right)-3\mathrm{cov}\left( \hat{b}_{f},\hat{b}_{p} \right)+\frac{3}{2}\mathrm{cov}\left( \hat{b}_{m},\hat{b}_{p} \right)}$$

$$=\sqrt{\frac{9}{4}\mathrm{var}\left( \hat{b}_{p} \right)+{\mathrm{var}(\hat{b}}_{f})+\frac{1}{4}\mathrm{var}\left( \hat{b}_{m} \right)-\mathrm{int}_{f,m}\times SE\left( \hat{b}_{f} \right)\mathrm{SE}(\hat{b}_{m})-3\times\mathrm{int}_{f,p}\times SE\left( \hat{b}_{f} \right)\mathrm{SE}(\hat{b}_{p})+\frac{3}{2}\times\mathrm{int}_{m,p}\times SE\left( \hat{b}_{m} \right)\mathrm{SE}(\hat{b}_{p})}$$

Likewise, the sampling covariance of fetal, maternal, paternal estimates is given by:

$${\mathrm{cov}(\hat{\beta}}_{f},\hat{\beta}_{m})=4\mathrm{cov}\left( \hat{b}_{f},\hat{b}_{m} \right)+2\mathrm{cov}\left( \hat{b}_{f},\hat{b}_{p} \right)-2\mathrm{cov}\left( \hat{b}_{m},\hat{b}_{p} \right)-2\mathrm{var}\left( \hat{b}_{f} \right)-\frac{3}{2}\mathrm{var}(\hat{b}_{m})-\frac{1}{2}\mathrm{var}\left( \hat{b}_{p} \right)$$

$$=4\times\mathrm{int}_{f,m}\times SE\left( \hat{b}_{f} \right)\mathrm{SE}(\hat{b}_{m})+2\times\mathrm{int}_{f,p}\times SE\left( \hat{b}_{f} \right)\mathrm{SE}(\hat{b}_{p})-2\times\mathrm{int}_{m,p}\times SE\left( \hat{b}_{m} \right)\mathrm{SE}(\hat{b}_{p})-2\mathrm{var}\left( \hat{b}_{f} \right)-\frac{3}{2}\mathrm{var}(\hat{b}_{m})-\frac{1}{2}\mathrm{var}\left( \hat{b}_{p} \right)$$

$${\mathrm{cov}(\hat{\beta}}_{f},\hat{\beta}_{p})=4\mathrm{cov}\left( \hat{b}_{f},\hat{b}_{p} \right)+2\mathrm{cov}\left( \hat{b}_{f},\hat{b}_{m} \right)-2\mathrm{cov}\left( \hat{b}_{m},\hat{b}_{p} \right)-2\mathrm{var}\left( \hat{b}_{f} \right)-\frac{3}{2}\mathrm{var}(\hat{b}_{p})-\frac{1}{2}\mathrm{var}\left( \hat{b}_{m} \right)$$

$$=4\times\mathrm{int}_{f,p}\times SE\left( \hat{b}_{f} \right)\mathrm{SE}(\hat{b}_{p})+2\times\mathrm{int}_{f,m}\times SE\left( \hat{b}_{f} \right)\mathrm{SE}(\hat{b}_{m})-2\times\mathrm{int}_{m,p}\times SE\left( \hat{b}_{m} \right)\mathrm{SE}(\hat{b}_{p})-2\mathrm{var}\left( \hat{b}_{f} \right)-\frac{3}{2}\mathrm{var}(\hat{b}_{p})-\frac{1}{2}\mathrm{var}\left( \hat{b}_{m} \right)$$

$${\mathrm{cov}(\hat{\beta}}_{m},\hat{\beta}_{p})=var\left( \hat{b}_{f} \right)+\frac{3}{4}\mathrm{var}\left( \hat{b}_{m} \right)+\frac{3}{4}\mathrm{var}\left( \hat{b}_{p} \right)+\frac{10}{4}\mathrm{cov}\left( \hat{b}_{m},\hat{b}_{p} \right)-2\mathrm{cov}(\hat{b}_{f},\hat{b}_{m})-2\mathrm{cov}\left( \hat{b}_{f},\hat{b}_{p} \right)$$

$$=var\left( \hat{b}_{f} \right)+\frac{3}{4}\mathrm{var}\left( \hat{b}_{m} \right)+\frac{3}{4}\mathrm{var}\left( \hat{b}_{p} \right)+\frac{10}{4}\times\mathrm{int}_{m,p}\times SE\left( \hat{b}_{m} \right)\mathrm{SE}(\hat{b}_{p})-2\times\mathrm{int}_{f,m}\times SE(\hat{b}_{f})\mathrm{SE}(\hat{b}_{m})-2\times\mathrm{int}_{f,p}\times SE\left( \hat{b}_{f} \right)\mathrm{SE}(\hat{b}_{p})$$

where int_f,m_, int_f,p_ and int_m,p_ are the estimated intercepts from the bivariate LD score regression of the maternal and fetal GWAS, the paternal and fetal GWAS, and the maternal and paternal GWAS respectively.

The joint sampling distribution of fetal, maternal and paternal effects is trivariate normal in large samples i.e.:

$$\left( \begin{matrix} {\hat{\beta}_{f} \atop\hat{\beta}_{m}} \\ \hat{\beta}_{p} \end{matrix} \right)\sim N\left( \binom{\beta_{f}}{\begin{matrix} \beta_{m} \\ \beta_{p} \end{matrix}} , \boldsymbol{\Sigma} \right)$$

with $\boldsymbol{\Sigma}=\left( \begin{matrix} \mathrm{var}(\hat{\beta}_{f}) & \mathrm{cov}(\hat{\beta}_{f},\hat{\beta}_{m}) & \mathrm{cov}(\hat{\beta}_{f},\hat{\beta}_{p}) \\ \mathrm{cov}(\hat{\beta}_{f},\hat{\beta}_{m}) & \mathrm{var}(\hat{\beta}_{m}) & \mathrm{cov}(\hat{\beta}_{m},\hat{\beta}_{p}) \\ \mathrm{cov}(\hat{\beta}_{f},\hat{\beta}_{p}) & \mathrm{cov}(\hat{\beta}_{m},\hat{\beta}_{p}) & \mathrm{var}(\hat{\beta}_{p}) \end{matrix} \right)$

We propose a test that considers the joint distribution of the three effect estimates. Using similar logic to above, it follows that under the null hypothesis ($H_{0}$) of no association between trait and maternal, paternal and fetal genotype, the test statistic:

$$\left( \begin{matrix} \hat{\beta}_{f} & \hat{\beta}_{m} & \hat{\beta}_{p} \end{matrix} \right)\left( \begin{matrix} \mathrm{var}(\hat{\beta}_{f}) & \mathrm{cov}(\hat{\beta}_{f},\hat{\beta}_{m}) & \mathrm{cov}(\hat{\beta}_{f},\hat{\beta}_{p}) \\ \mathrm{cov}(\hat{\beta}_{f},\hat{\beta}_{m}) & \mathrm{var}(\hat{\beta}_{m}) & \mathrm{cov}(\hat{\beta}_{m},\hat{\beta}_{p}) \\ \mathrm{cov}(\hat{\beta}_{f},\hat{\beta}_{p}) & \mathrm{cov}(\hat{\beta}_{m},\hat{\beta}_{p}) & \mathrm{var}(\hat{\beta}_{p}) \end{matrix} \right)^{-1}\left( \begin{matrix} {\hat{\beta}_{f} \atop\hat{\beta}_{m}} \\ \hat{\beta}_{p} \end{matrix} \right)\sim X_{3}^{2}$$

is distributed as a central chi-square statistic with three degrees of freedom.

***Formulation of One Degree of Freedom Meta-analytic Tests when Fathers’ GWAS Available***

It is straight forward to extend the meta-analytic test for fetal effects to include information from a GWAS of fathers i.e.:

$$w_{1}=\frac{1}{\mathrm{var}(\hat{b}_{f})}$$

$$w_{2}=\frac{1}{4var(\hat{b}_{m})}$$

$$w_{3}=\frac{1}{4var(\hat{b}_{p})}$$

$$\hat{\beta}_{f\_meta}=\frac{{w_{1}\hat{b}}_{f}+{{2w}_{2}\hat{b}}_{m}+{{2w}_{3}\hat{b}}_{p}}{w_{1}+w_{2}+w_{3}}$$

$${\mathrm{SE}(\hat{\beta}}_{f\_meta})=\sqrt{\begin{aligned} (\frac{w_{1}}{w_{1}+w_{2}+w_{3}})^{2}\mathrm{var}(\hat{b}_{f})+4(\frac{w_{2}}{w_{1}+w_{2}+w_{3}})^{2}\mathrm{var}\left( \hat{b}_{m} \right)+4(\frac{w_{3}}{w_{1}+w_{2}+w_{3}})^{2}\mathrm{var}\left( \hat{b}_{p} \right)+ \\ 4\mathrm{cov}\left( \hat{b}_{f},\hat{b}_{m} \right)\frac{{w_{1}w}_{2}}{\left( w_{1}+w_{2}+w_{3} \right)^{2}}+4\mathrm{cov}\left( \hat{b}_{f},\hat{b}_{p} \right)\frac{{w_{1}w}_{3}}{\left( w_{1}+w_{2}+w_{3} \right)^{2}}+8\mathrm{cov}(\hat{b}_{m},\hat{b}_{p})\frac{{w_{2}w}_{3}}{{(w_{1}+w_{2}+w_{3})}^{2}} \end{aligned}}$$

where $\hat{b}_{f}$ is the coefficient from the regression of own phenotype on own genotype, $\hat{b}_{m}$ is the coefficient from the regression of offspring phenotype on maternal genotype, $\hat{b}_{p}$ is the coefficient from the regression of offspring phenotype on paternal genotype, and $\hat{\beta}_{f\_meta}$ the inverse variance weighted estimate of the fetal effect across all three scans. The effective sample overlap between the scans is estimated using bivariate LD score regression and the covariance between the regression coefficients estimated from this quantity:

$$\hat{\mathrm{cov}}\left( \hat{b}_{f},\hat{b}_{m} \right)\approx\frac{N_{S\_fm}}{\sqrt{N_{f}N_{m}}}\rho_{fm}\sqrt{{\mathrm{var}(\hat{b}}_{f})\mathrm{var}(\hat{b}_{m})}$$

$$=\hat{int}_{fm}\times{\mathrm{SE}(\hat{b}}_{f})\mathrm{SE}(\hat{b}_{m})$$

$$\hat{\mathrm{cov}}\left( \hat{b}_{f},\hat{b}_{p} \right)\approx\frac{N_{S\_fp}}{\sqrt{N_{f}N_{p}}}\rho_{fp}\sqrt{{\mathrm{var}(\hat{b}}_{f})\mathrm{var}(\hat{b}_{p})}$$

$$=\hat{int}_{fp}\times{\mathrm{SE}(\hat{b}}_{f})\mathrm{SE}(\hat{b}_{p})$$

$$\hat{\mathrm{cov}}\left( \hat{b}_{m},\hat{b}_{p} \right)\approx\frac{N_{S\_mp}}{\sqrt{N_{m}N_{p}}}\rho_{mp}\sqrt{{\mathrm{var}(\hat{b}}_{m})\mathrm{var}(\hat{b}_{p})}$$

$$=\hat{int}_{mp}\times{\mathrm{SE}(\hat{b}}_{m})\mathrm{SE}(\hat{b}_{p})$$

where $N_{f}$ is the sample size of the GWAS of own genotype and own outcome, $N_{m}$ is the sample size of the GWAS of maternal genotype and offspring phenotype, $N_{p}$ is the sample size of the GWAS of paternal genotype and offspring phenotype, $N_{S\_fm}$, $N_{S\_fp}$, and $N_{S\_mp}$ are the effective number of overlapping individuals across the relevant GWAS, and $\hat{int}_{fm}$, $\hat{int}_{fp}$, and $\hat{int}_{mp}$ are the associated estimated bivariate LD score regression intercepts. The parameters $\rho_{fm}$, $\rho_{fp}$, and $\rho_{mp}$ refers to the correlation between the phenotype in the relevant overlapping individuals.

We note that the effective sample overlap between maternal and paternal scans is likely to be small but could still be non-zero (e.g. for example if brothers and their sisters were in the fathers’ and mothers’ GWAS respectively).

***Power of the one degree of freedom unconditional fetal and maternal test of association***

Without loss of generality, we assume that the SNP marker and the outcome have been standardized to unit variance. The sampling variance of the (biased) ordinary least squares estimate of the fetal genetic effect is:

$$\mathrm{var}(\hat{b}_{f})=\frac{(1-{{(\beta}_{f}+\frac{1}{2}\beta_{m})}^{2})}{N_{f}}$$

and likewise for (biased) ordinary least squares estimate of the maternal genetic effect:

$$\mathrm{var}(\hat{b}_{m})=\frac{(1-{{(\beta}_{m}+\frac{1}{2}\beta_{f})}^{2})}{N_{m}}$$

where *N_f_* and *N_m_* are the number of individuals in the fetal and maternal GWAS respectively.

It follows that the non-centrality parameter (NCP) for the unconditional one degree of freedom tests for fetal and maternal effects are given by:

$${NCP}_{FETAL}=\frac{{(\beta}_{f}+\frac{1}{2}\beta_{m})^{2}}{\mathrm{var}(\hat{b}_{f})}$$

$${NCP}_{MATERNAL}=\frac{{(\beta}_{m}+\frac{1}{2}\beta_{f})^{2}}{\mathrm{var}(\hat{b}_{m})}$$

Asymptotic power can then be calculated as the area under the curve of a non-central chi-square distribution to the right of the significance threshold of interest:

$$Power= \int_{X_{\alpha}^{'2}(v,0)}^{\infty} d{X^{'}}^{2}\left( v,\mathrm{NCP} \right)$$

where $X_{\alpha}^{'2}(v,0)$ is the quantile of the 100 * (1-α) percentage point of the central *χ^2^* distribution with $v$ degrees of freedom (one in this case), and NCP is the non-centrality parameter.

In order to estimate the power that either the maternal or fetal effect is significant we sample standardized effect sizes under the alternative hypothesis from a bivariate normal distribution with mean vector:

$$\boldsymbol{\mu}=\begin{matrix} \frac{\beta_{f}+\frac{1}{2}\beta_{m}}{\sqrt{\mathrm{var}(\hat{b}_{f})}} \\ \frac{\beta_{m}+\frac{1}{2}\beta_{f}}{\sqrt{\mathrm{var}(\hat{b}_{m})}} \end{matrix}$$

and covariance matrix:

$$\boldsymbol{\Sigma}=\begin{matrix} 1 & \frac{N_{S}}{\sqrt{N_{f}N_{m}}}\rho\sqrt{{\mathrm{var}(\hat{b}}_{f})\mathrm{var}(\hat{b}_{m})} \\ \frac{N_{S}}{\sqrt{N_{f}N_{m}}}\rho\sqrt{{\mathrm{var}(\hat{b}}_{f})\mathrm{var}(\hat{b}_{m})} & 1 \end{matrix}$$

Power is calculated as the proportion of draws where the absolute fetal *or* maternal effect exceeds the relevant critical value from the univariate standard normal distribution.

***Power of the one degree of freedom meta-analytic test for fetal and maternal effects***

Recall that the weights for the meta-analysis of fetal genetic effects are given by:

$$w_{1}=\frac{1}{\mathrm{var}(\hat{b}_{f})}$$

$$w_{2}=\frac{1}{4var(\hat{b}_{m})}$$

leading to the weighted estimate:

$$\hat{\beta}_{f\_meta}=\frac{w_{1}\hat{b}_{f}+{2w_{2}\hat{b}}_{m}}{w_{1}+w_{2}}$$

and its standard error:

$${\mathrm{SE}(\hat{\beta}}_{f\_meta})=\sqrt{(\frac{w_{1}}{w_{1}+w_{2}})^{2}\mathrm{var}(\hat{b}_{f})+4(\frac{w_{2}}{w_{1}+w_{2}})^{2}\mathrm{var}\left( \hat{b}_{m} \right)+4\mathrm{cov}(\hat{b}_{f},\hat{b}_{m})\frac{{w_{1}w}_{2}}{{{(w}_{1}+w_{2})}^{2}}}$$

Recall that the expectation for the fetal and maternal regression coefficients are given by:

$E(\hat{b}_{f})=$ $\beta_{f}+\beta_{m}\frac{\mathrm{cov}(x_{f},x_{m})}{\mathrm{var}(x_{f})}= \beta_{f}+{\frac{1}{2}\beta}_{m}$ in large samples,

$E(\hat{b}_{m})=$ $\beta_{m}+\beta_{f}\frac{\mathrm{cov}(x_{f},x_{m})}{\mathrm{var}(x_{m})}= \beta_{m}+{\frac{1}{2}\beta}_{f}$ in large samples

It follows that the non-centrality parameter for the meta-analytic fetal test is given by:

$${NCP}_{FETAL\_META}=\frac{{(\frac{w_{1}(\beta_{f}+{\frac{1}{2}\beta}_{m})+{2w_{2}(\beta}_{m}+{\frac{1}{2}\beta}_{f})}{w_{1}+w_{2}})}^{2}}{{\mathrm{SE}(\hat{\beta}_{f\_meta})}^{2}}$$

The weights for the meta-analysis of maternal genetic effects are given by:

$$w_{3}=\frac{1}{4var(\hat{b}_{f})}$$

$$w_{4}=\frac{1}{\mathrm{var}(\hat{b}_{m})}$$

leading to the weighted estimate:

$$\hat{\beta}_{f\_meta}=\frac{{{2w}_{3}\hat{b}}_{f}+{w_{4}\hat{b}}_{m}}{w_{3}+w_{4}}$$

and its standard error

$${\mathrm{SE}(\hat{\beta}}_{m\_meta})=\sqrt{(\frac{w_{3}}{w_{3}+w_{4}})^{2}\mathrm{var}(\hat{b}_{m})+4(\frac{w_{3}}{w_{3}+w_{4}})^{2}\mathrm{var}\left( \hat{b}_{f} \right)+4\mathrm{cov}(\hat{b}_{f},\hat{b}_{m})\frac{{w_{3}w}_{4}}{{{(w}_{3}+w_{4})}^{2}}}$$

It follows that the non-centrality parameter for the meta-analytic maternal test is given by:

$${NCP}_{MATERNAL\_META}=\frac{{(\frac{{2w}_{3}(\beta_{f}+{\frac{1}{2}\beta}_{m})+{w_{4}(\beta}_{m}+{\frac{1}{2}\beta}_{f})}{w_{3}+w_{4}})}^{2}}{{\mathrm{SE}(\hat{\beta}_{m\_meta})}^{2}}$$

In order to estimate the power that either the maternal or fetal meta-analytic effect is significant we sample standardized effect sizes under the alternative hypothesis from a bivariate normal distribution with mean:

$$\boldsymbol{\mu}=\begin{matrix} \frac{\frac{w_{1}(\beta_{f}+{\frac{1}{2}\beta}_{m})+{2w_{2}(\beta}_{m}+{\frac{1}{2}\beta}_{f})}{w_{1}+w_{2}}}{{\mathrm{SE}(\hat{\beta}}_{f\_meta})} \\ \frac{\frac{{2w}_{3}(\beta_{f}+{\frac{1}{2}\beta}_{m})+{w_{4}(\beta}_{m}+{\frac{1}{2}\beta}_{f})}{w_{3}+w_{4}}}{{\mathrm{SE}(\hat{\beta}}_{m\_meta})} \end{matrix}$$

and correlation matrix:

$$\boldsymbol{\Sigma}=\begin{matrix} 1 & \mathrm{cor}(\hat{\beta}_{f_{meta}},\hat{\beta}_{m\_meta}) \\ \mathrm{cor}(\hat{\beta}_{f_{meta}},\hat{\beta}_{m\_meta}) & 1 \end{matrix}$$

where

$\mathrm{cor}\left( \hat{\beta}_{f_{meta}},\hat{\beta}_{m_{meta}} \right)=\left( \left( \frac{w_{1}}{w_{1}+w_{2}} \right)\left( \frac{{2w}_{3}}{w_{3}+w_{4}} \right)\mathrm{var}\left( \hat{b}_{f} \right)+\left( \frac{w_{4}}{w_{3}+w_{4}} \right)\left( \frac{{2w}_{2}}{w_{1}+w_{2}} \right)\mathrm{var}\left( \hat{b}_{m} \right)+\left( \left( \frac{w_{1}}{w_{1}+w_{2}} \right)\left( \frac{w_{4}}{w_{3}+w_{4}} \right)+\left( \frac{{2w}_{2}}{w_{1}+w_{2}} \right)\left( \frac{{2w}_{3}}{w_{3}+w_{4}} \right) \right)\mathrm{cov}\left( \hat{b}_{m},\hat{b}_{f} \right) \right)/$(${\mathrm{SE}(\hat{\beta}}_{f\_meta}){\mathrm{SE}(\hat{\beta}}_{m\_meta})$)

Power is calculated as the proportion of draws where the absolute fetal *or* maternal effect exceeds the relevant critical value from the univariate standard normal distribution.

**Distribution of the maximum chi-square statistic from the fetal and maternal GWAS under the null hypothesis of no association and derivation of the correction for multiple testing**

The one degree of freedom strategies investigated in this manuscript involve double the number of statistical tests across the genome as the two degree of freedom strategy. We address this issue by dividing the significance threshold for the one degree of freedom strategies by two (i.e. adjusting the threshold for genome-wide significance from α = 6.6 x 10^-9^ to α = 3.3 x 10^-9^). To investigate the appropriateness of this procedure, we simulated the distribution of the maximum chi-square statistic (i.e. from potentially correlated fetal and maternal tests of association) under the null hypothesis of no association (1 million draws) whilst varying the sample overlap (we assume 1000 individuals in the maternal test, and 1000 individuals in the fetal test) and the correlation between maternal and fetal phenotype. We calculate the quantile corresponding to the upper 5%, 1% and 0.1% tail of this maximum chi-square distribution. We then determine what upper tail probability of the one degree of freedom central chi-square distribution this value corresponds to (Supplementary Table 1). Our results suggest, that even in the case of 100% overlap and ρ = 0.5, a Bonferroni correction is not too conservative.

**Supplementary Table 1** Upper tail probabilities of the one degree of freedom central chi-square distribution associated with the corresponding quantile from the maximum chi-square distribution.

|  | α = 0.05 | α = 0.01 | α = 0.001 |
| --- | --- | --- | --- |
| 0% overlap, ρ = 0 | 0.0253 | 0.00500 | 0.000496 |
| 50% overlap, ρ = 0 | 0.0251 | 0.00503 | 0.000485 |
| 100% overlap, ρ = 0 | 0.0252 | 0.00505 | 0.000484 |
| 0% overlap, ρ = 0.5 | 0.0253 | 0.00498 | 0.000487 |
| 50% overlap, ρ = 0.5 | 0.0256 | 0.00503 | 0.000501 |
| 100% overlap, ρ = 0.5 | 0.0269 | 0.00525 | 0.000536 |
